# Leveraging Unstructured Data in Electronic Health Records to Detect Adverse Events from Pediatric Drug Use - A Scoping Review

**DOI:** 10.1101/2025.03.20.25324320

**Authors:** Su Golder, Karen O’Connor, Guillermo Lopez-Garcia, Nicholas Tatonetti, Graciela Gonzalez-Hernandez

## Abstract

Adverse drug events (ADEs) in pediatric populations pose significant public health challenges, yet research on their detection and monitoring remains limited. This scoping review evaluates the use of unstructured data from electronic health records (EHRs) to identify ADEs in children. We searched six databases, including MEDLINE, Embase and IEEE Xplore, in September 2024. From 984 records, only nine studies met our inclusion criteria, indicating a significant gap in research towards identify ADEs in children. We found that unstructured data in EHRs can indeed be of value and enhance pediatric pharmacovigilance, although its use has been so far very limited. Traditional Natural Language Processing (NLP) methods have been employed to extract ADEs, but the approaches utilized face challenges in generalizability and context interpretation. These challenges could be addressed with recent advances in transformer-based models and large language models (LLMs), unlocking the use of EHR data at scale for pediatric pharmacovigilance.

## Introduction

Adverse drug events (ADEs) represent a significant challenge in public health (1, 2). Even seemingly minor ADEs can profoundly affect a patient’s quality of life and treatment adherence, depriving patients of potentially beneficial treatments (3). The effect of ADEs is particularly acute in pediatric populations(4), where the prevalence of ADEs can reach as high as 16.8% of all children exposed to a drug during hospital stay (5).

Children’s unique physiological characteristics (4, 6), coupled with the scarcity of pediatric-specific drug safety and efficacy data, make them especially vulnerable to ADEs (7). The frequent use of off-label and unlicensed prescribing further compounds this risk (8–11), leading to more hospital admissions resulting from ADEs in children than in adults (12–14). ADEs account for up to 10% of pediatric hospitalizations (5), with up to 45% of them being life threatening (15). Moreover, the overall incidence of ADEs during hospital stays is also reported around 10% (15, 16) and is similarly high in children attending outpatient clinics (5), where many cases are likely to go unreported.

Despite the extensive literature on ADEs in adult populations, relatively little is known about their frequency and nature in pediatric populations (17, 18). This knowledge gap stems from various challenges inherent in pediatric clinical trials, including ethical concerns, recruitment difficulties, and a lack of established endpoints (19). Consequently, much of the current knowledge about drug effects in children is extrapolated from adult studies (6, 20–22), despite known differences in drug responses between these populations (23).

In the absence of comprehensive pediatric clinical trial data, post marketing surveillance becomes crucial for monitoring drug safety in children. Traditional pharmacovigilance systems, relying on spontaneous reporting to regulatory agencies, have significant limitations, including underreporting and incomplete data (24–26). This has prompted researchers to look at new methods of mining regulatory data for pediatric drug safety signals (27, 28). These shortcomings have also spurred interest in alternative data sources and methodologies for detecting drug safety signals (29).

Electronic health records (EHRs) have emerged as a promising resource for pharmacovigilance, offering detailed longitudinal and demographic data. The vast amount of unstructured data within EHRs, including clinical notes and discharge summaries, can provide valuable insights and potentially aide pharmacovigilance. However, extracting knowledge from unstructured data requires specialized Natural Language Processing (NLP) approaches that are not readily available.

Routine electronic healthcare records have been used for pharmacovigilance in children (30). However, most existing studies focus on structured data (30–32) or extrapolate from adult populations (33–35). This review provides an overview of studies using unstructured EHR data in ADE detecting and reflects on how it can be used to enhance pharmacovigilance in pediatric populations.

## Methods

We conducted a scoping review following the methodology outlined by Arksey and O’Malley (36) to explore recent research on detecting ADEs in children using unstructured data from EHRs/EMRs. This scoping review adheres to the PRISMA extension for Scoping Reviews (PRISMA-ScR) guidelines for reporting a systematic and transparent approach (37). A protocol was written by the lead author and agreed by the team.

### Inclusion and exclusion criteria

To comprehensively assess the volume and nature of research in this area, strict inclusion and exclusion criteria were established (Table 1). Studies were considered eligible if they used any type of unstructured EHRs/EMRs (such as clinical notes or discharge summaries) to detect any type of adverse event associated with any type of medication in children (defined as <18 years). This approach allowed us to capture a broad range of relevant studies while maintaining a focused scope on the use of unstructured EHR/EMR data for pediatric pharmacovigilance.

**Table 1.** Inclusion and exclusion criteria for studies on identifying adverse drug events data in Children from EHRs.

|  | Inclusion criteria | Exclusion criteria |
| --- | --- | --- |
| **Population** | Children aged <18 years old. This included studies with subgroup analysis for pediatrics) | Adults aged ≥18 years old |
| **Intervention(s)** | Extraction of adverse events documented in unstructured text in clinical data such as EHRs/EMRs or administrative data (typically using NLP/ML/LLM) | Studies focused on structured health records or structured administrative data |
| **Comparator(s)** | No comparator was required | No study was excluded based on the comparator |
| **Outcomes:** | Primary outcome: The feasibility of using unstructured text in the identification of pediatrics adverse events. | We were concerned with the properties of interventions under normal use. We therefore did not consider papers where the primary aim was to assess events such as intentional and accidental poisoning (i.e., overdose), drug abuse, errors, or non-compliance.<br>Drug–drug interactions were also not eligible where they were the primary objective of the paper due to the different techniques required in identifying interactions as opposed to adverse events under normal use. |
| **Setting** | Any healthcare setting from any geographical location. | Non-healthcare settings. |
| **Study design:** | Any type of assessment. | Non-empirical research, opinion pieces, commentaries, letters to the editors, or editorials. |
| **Limits** | Published 2000 onwards in English (or translation available). | Published before 2000 or non-English language. |

### Search Methods

A comprehensive literature search was conducted across six databases covering a range of topic areas, including health and medical sciences (MEDLINE and Embase), psychology (PsycINFO), and information and computer science (Library, Information Science & Technology Abstracts (LISTA) database, ACM Digital Library, and IEEE Xplore). The database search strategies incorporated four key facets which were combined using the Boolean AND operator. For each of the four facets namely – “Natural Language Processing”, “Electronic Health Records”, “Adverse Events” and “Pediatrics” multiple synonyms and indexing terms were combined with the Boolean OR operator (see supplementary material for full strategies). Thus the final set of records contained at least one term from each of the four facets. A publication date restriction of 2000 onwards was placed on the searches as automative methods before this date will not be applicable to current methods. No language restrictions were applied, although financial and logistical restraints did not allow translation from all languages.

To enhance the comprehensiveness of the search, forward and backward citation searching was performed on the included studies using CitationChaser. All search results were imported into an Endnote library for deduplication. Title and abstract screening was undertaken independently by two reviewers in Covidence (ref) with any disagreements resolved by discussion or, if necessary, a third reviewer. Full-text screening was again undertaken in Covidence by two independent reviewers.

### Data Extraction

A customized data extraction form was developed and tested for this review in Covidence to capture key information from the included studies. The form recorded the study characteristics of existing papers on using unstructured EMR data to identify potential adverse drug events in children. Two reviewers extracted the descriptive data independently, with findings compared and agreed. The following data was extracted from included studies if available:

1. Details on the type of data used.
2. Details on the age of the children studied.
3. Details on the primary aim of the study.
4. Brief details of the methods used to extract data from unstructured EMR including which drugs or adverse events are searched for.
5. The type and frequency of adverse events data identified for each drug and which drug.
6. Conclusions of the original investigators.
7. Lastly, whether code or annotated or raw data are made available by the authors.

As this is a scoping review, we did not assess the methodological quality (risk of bias assessment) of the studies or conduct any evidence synthesis. Nevertheless, we do summarise the array of Artificial Intelligence (AI), Natural Language Processing (NLP), and Machine Learning (ML) methods used for this task and synthesize the studies’ self-reported performances and if available, scalability per method.

### Ethical Considerations

Since the scoping review methodology consists of reviewing and collecting data from publicly accessible materials, this study does not require ethical approval.

## Results

A total of 984 records were identified by the searches and citation searches, 833 of which were unique records. 90 full-text articles were obtained after screening the title and abstract and only 9 studies met our inclusion criteria (Figure 1: Flow diagram for included studies). The 81 excluded studies are listed in supplementary materials. The most common reasons for excluding papers at the full-text stage were: they did not use unstructured data (often limited to structured data), they were not limited to a pediatric population or did not have results presented separately for pediatrics, or they were not studying a drug intervention.

**Figure 1:**
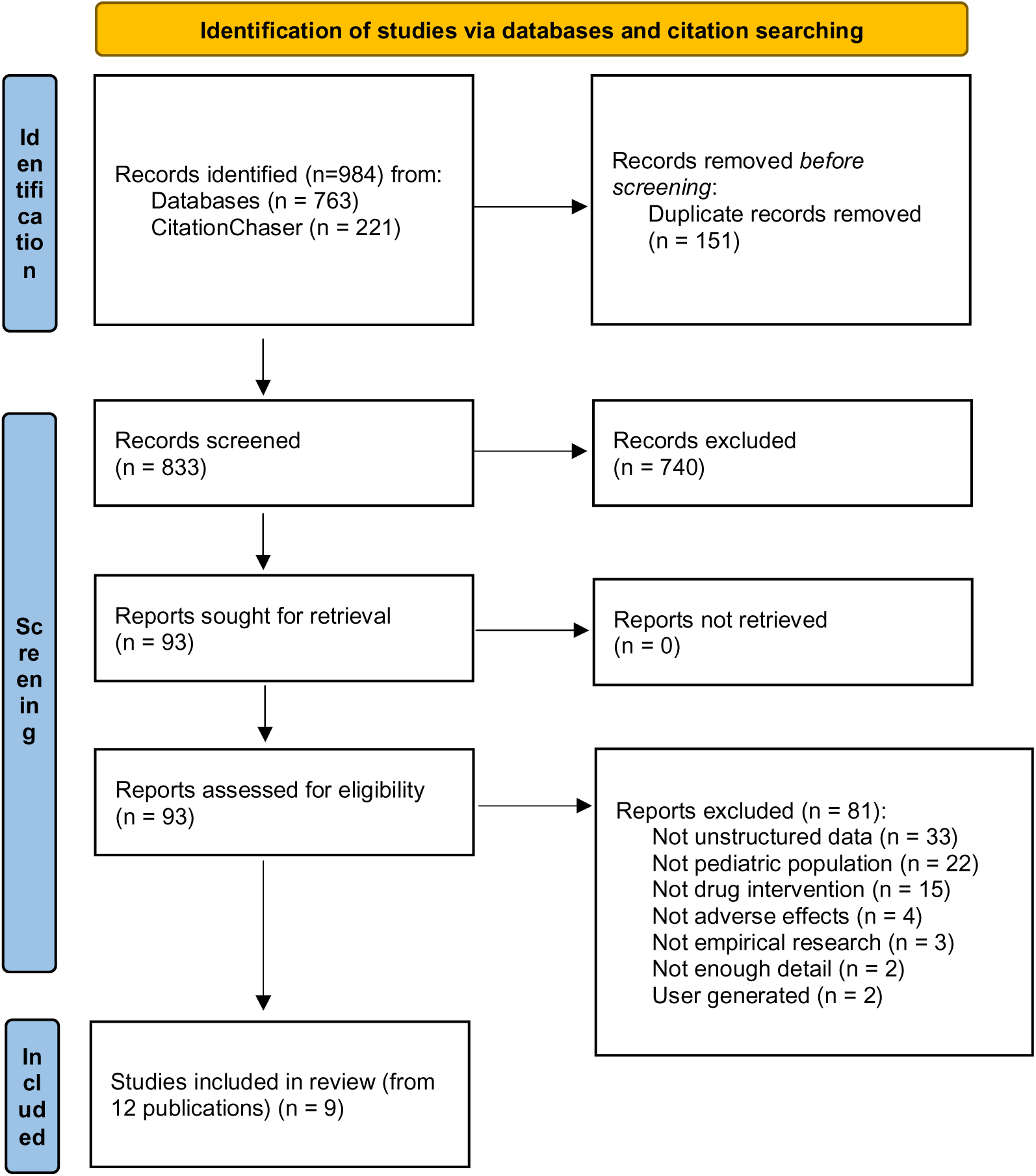
Flow diagram for included studies

### Characteristics of Included Studies

The nine included studies were published between 2017 and 2022 (Table 2). Four of the nine included studies used the same dataset as another study, with Aldrich 2019 (38) and Ramsey 2019 (39) using one dataset and Geva 2020 (40) using the same initial data as Geva 2019 (41). The majority of the studies were conducted in the USA with only one vanderStoep 2021A and B (42, 43) conducted in the Netherlands. The population sizes varied greatly from 206 patients in vanderStoep 2021A and B (42, 43) to 56,436 in Matson 2023 (44) with one study not reporting the number of patients - Zheng 2022A and B (45, 46). While all studies focused on children and adolescents, one study, Miller 2022 (47, 48), extended their age range to 22 years. Gender distribution was reported in some studies, with a relatively even split between males and females in most cases. Race and ethnicity were reported in some studies, with white patients often being the majority. All studies selected drug-ADE pairs based on a priori knowledge. The drugs studied ranged from specific medications (es/citalopram, treosulfan, bulsulfan, sildenafil, tadalafil, bosentan, ambrisentan) to broader categories such as chemotherapy, vaccines, and the most frequently used medications. Some studies focused on one specific ADE (such as weight gain, shoulder injury or myalgia), whilst others include a diverse range of ADEs.

**Table 2:**
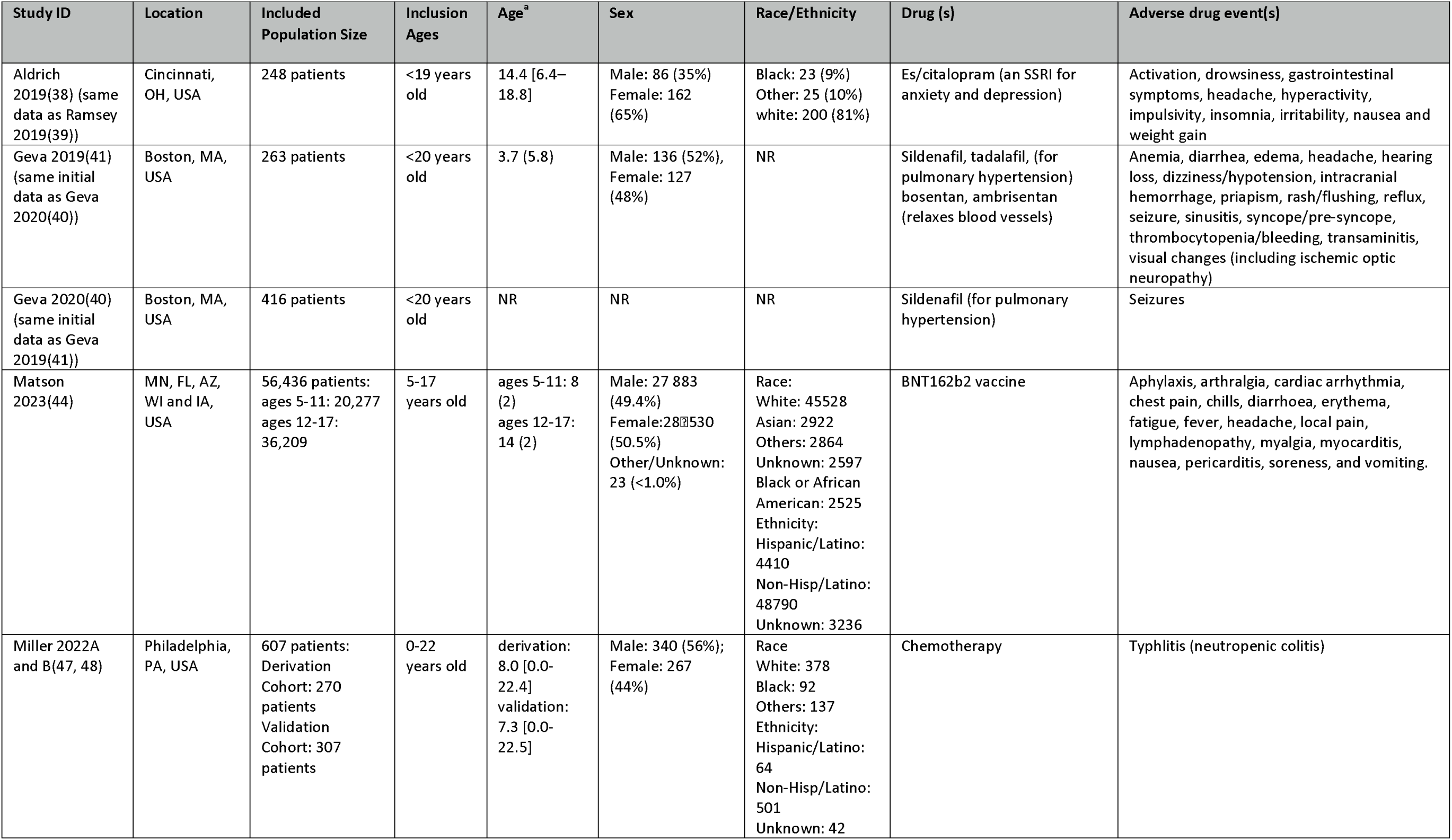
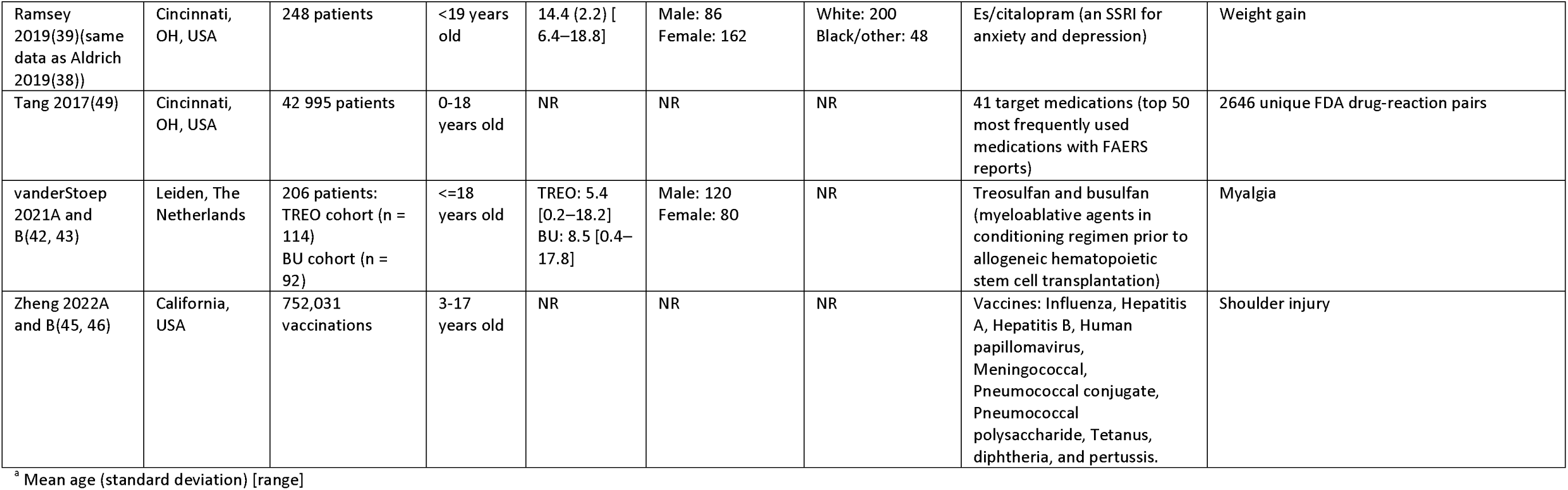
Characteristics of the included studies.

| Study ID | Location | Included Population Size | Inclusion Ages | Age<sup>a</sup> | Sex | Race/Ethnicity | Drug (s) | Adverse drug event(s) |
| --- | --- | --- | --- | --- | --- | --- | --- | --- |
| Aldrich 2019(38) (same data as Ramsey 2019(39)) | Cincinnati, OH, USA | 248 patients | <19 years old | 14.4 [6.4–18.8] | Male: 86 (35%)<br>Female: 162 (65%) | Black: 23 (9%)<br>Other: 25 (10%)<br>white: 200 (81%) | Es/citalopram (an SSRI for anxiety and depression) | Activation, drowsiness, gastrointestinal symptoms, headache, hyperactivity, impulsivity, insomnia, irritability, nausea and weight gain |
| Geva 2019(41) (same initial data as Geva 2020(40)) | Boston, MA, USA | 263 patients | <20 years old | 3.7 (5.8) | Male: 136 (52%),<br>Female: 127 (48%) | NR | Sildenafil, tadalafil, (for pulmonary hypertension) bosentan, ambrisentan (relaxes blood vessels) | Anemia, diarrhea, edema, headache, hearing loss, dizziness/hypotension, intracranial hemorrhage, priapism, rash/flushing, reflux, seizure, sinusitis, syncope/pre-syncope, thrombocytopenia/bleeding, transaminitis, visual changes (including ischemic optic neuropathy) |
| Geva 2020(40) (same initial data as Geva 2019(41)) | Boston, MA, USA | 416 patients | <20 years old | NR | NR | NR | Sildenafil (for pulmonary hypertension) | Seizures |
| Matson 2023(44) | MN, FL, AZ, WI and IA, USA | 56,436 patients:<br>ages 5-11: 20,277<br>ages 12-17: 36,209 | 5-17 years old | ages 5-11: 8 (2)<br>ages 12-17: 14 (2) | Male: 27 883 (49.4%)<br>Female: 28 530 (50.5%)<br>Other/Unknown: 23 (<1.0%) | Race:<br>White: 45528<br>Asian: 2922<br>Others: 2864<br>Unknown: 2597<br>Black or African American: 2525<br>Ethnicity:<br>Hispanic/Latino: 4410<br>Non-Hisp/Latino: 48790<br>Unknown: 3236 | BNT162b2 vaccine | Aphylaxis, arthralgia, cardiac arrhythmia, chest pain, chills, diarrhoea, erythema, fatigue, fever, headache, local pain, lymphadenopathy, myalgia, myocarditis, nausea, pericarditis, soreness, and vomiting. |
| Miller 2022A and B(47, 48) | Philadelphia, PA, USA | 607 patients:<br>Derivation Cohort: 270 patients<br>Validation Cohort: 307 patients | 0-22 years old | derivation: 8.0 [0.0-22.4]<br>validation: 7.3 [0.0-22.5] | Male: 340 (56%);<br>Female: 267 (44%) | Race<br>White: 378<br>Black: 92<br>Others: 137<br>Ethnicity:<br>Hispanic/Latino: 64<br>Non-Hisp/Latino: 501<br>Unknown: 42 | Chemotherapy | Typhlitis (neutropenic colitis) |
| Ramsey 2019(39)(same data as Aldrich 2019(38)) | Cincinnati, OH, USA | 248 patients | <19 years old | 14.4 (2.2) [6.4–18.8] | Male: 86<br>Female: 162 | White: 200<br>Black/other: 48 | Es/citalopram (an SSRI for anxiety and depression) | Weight gain |
| Tang 2017(49) | Cincinnati, OH, USA | 42 995 patients | 0-18 years old | NR | NR | NR | 41 target medications (top 50 most frequently used medications with FAERS reports) | 2646 unique FDA drug-reaction pairs |
| vanderStoep 2021A and B(42, 43) | Leiden, The Netherlands | 206 patients: TREO cohort (n = 114)<br>BU cohort (n = 92) | <=18 years old | TREO: 5.4 [0.2–18.2]<br>BU: 8.5 [0.4–17.8] | Male: 120<br>Female: 80 | NR | Treosulfan and busulfan (myeloablative agents in conditioning regimen prior to allogeneic hematopoietic stem cell transplantation) | Myalgia |
| Zheng 2022A and B(45, 46) | California, USA | 752,031 vaccinations | 3-17 years old | NR | NR | NR | Vaccines: Influenza, Hepatitis A, Hepatitis B, Human papillomavirus, Meningococcal, Pneumococcal conjugate, Pneumococcal polysaccharide, Tetanus, diphtheria, and pertussis. | Shoulder injury |

<sup>a</sup> Mean age (standard deviation) [range]

### Methods used in included studies

Different NLP techniques were used (Table 3). Aldrich 2019 (38), Ramsey 2019 (39), and Miller 2022A and B (47, 48) all used regular expressions or rule-based NLP algorithms, whereas Tang 2017 (49), Geva 2019 (41) and Geva 2020 (40) used hybrid approaches using (or basing their model on) a previously developed system, the Apache clinical Text Analysis Knowledge Extraction System (cTAKES) which combines rule-based learning and machine learning techniques. Matson 2023 used transformer-based classification (44) and other word embedding methods were used by Zheng 2022 (45, 46). One study used commercial software to mine the clinical text for symptom mentions - vanderStoep 2021 (42, 43).

**Table 3:**
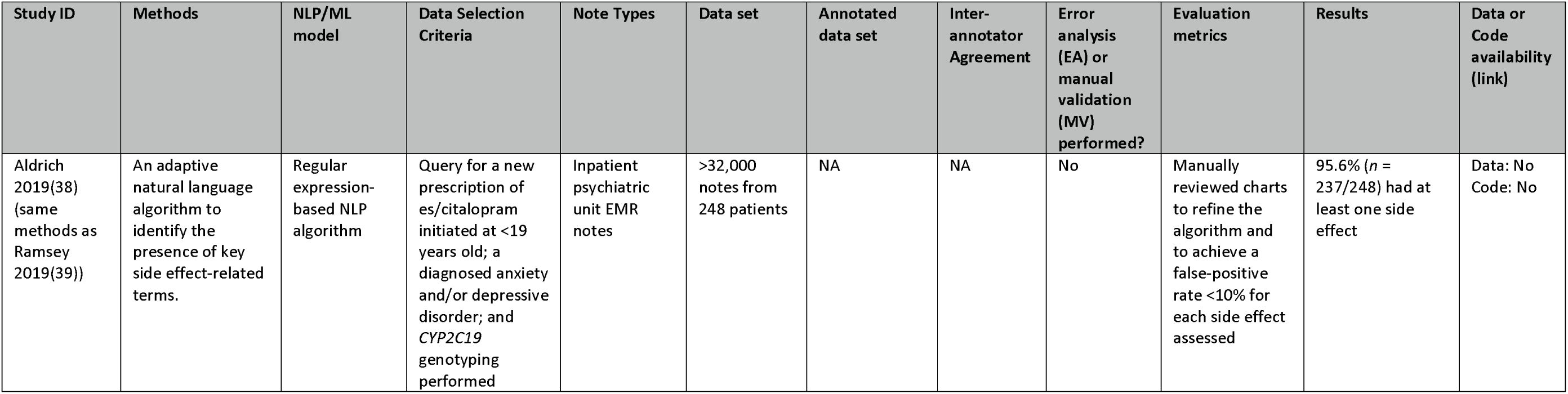
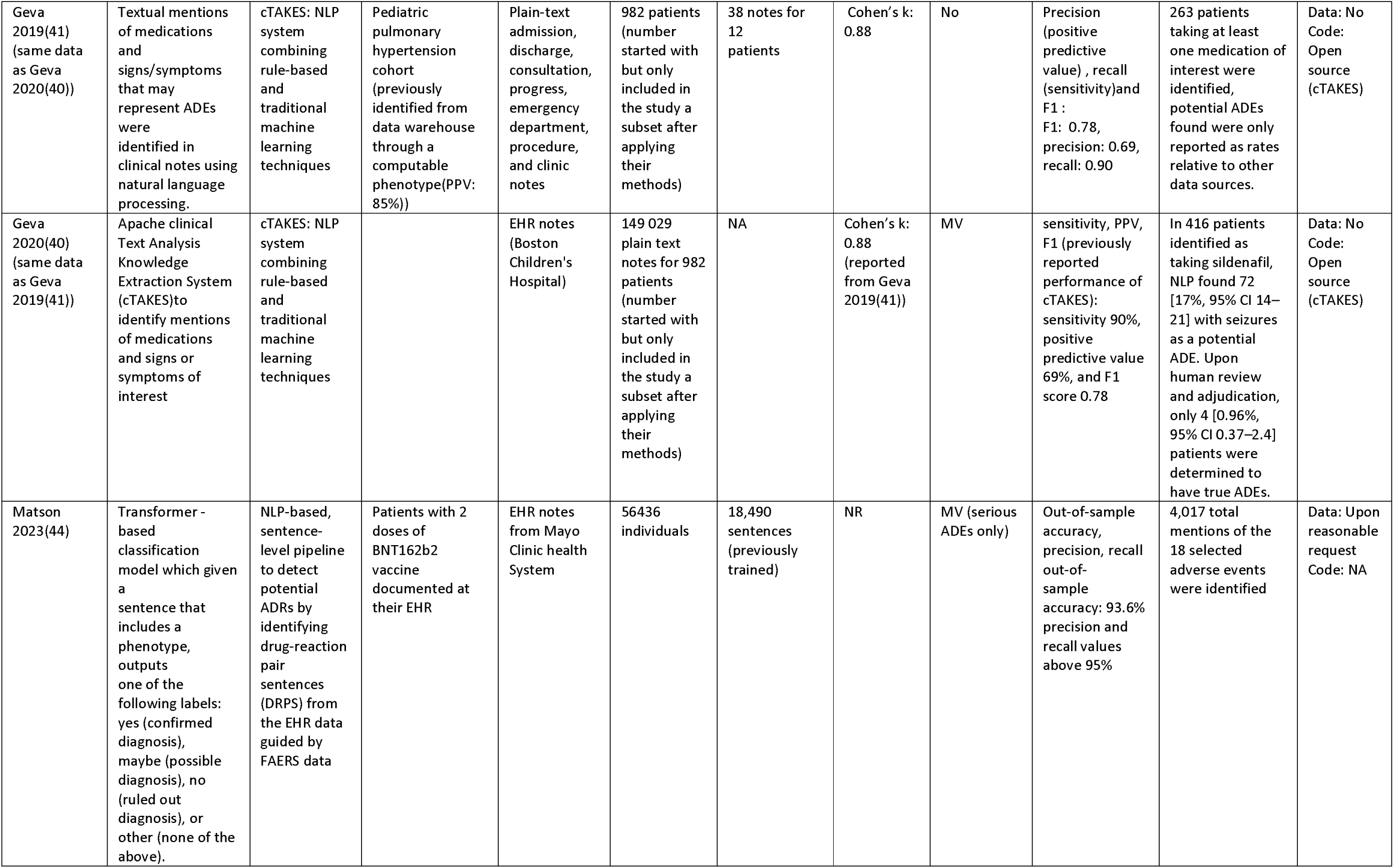
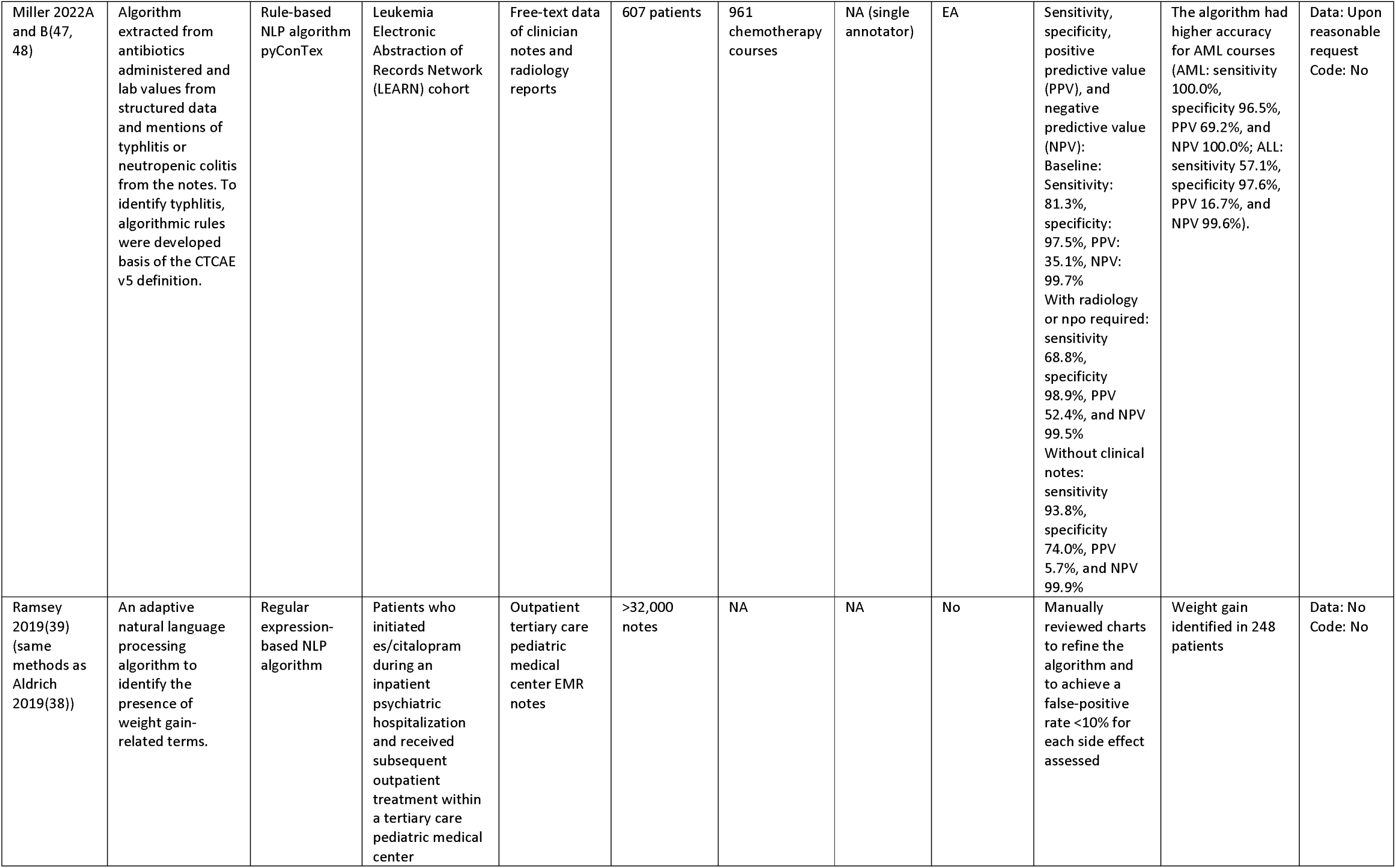
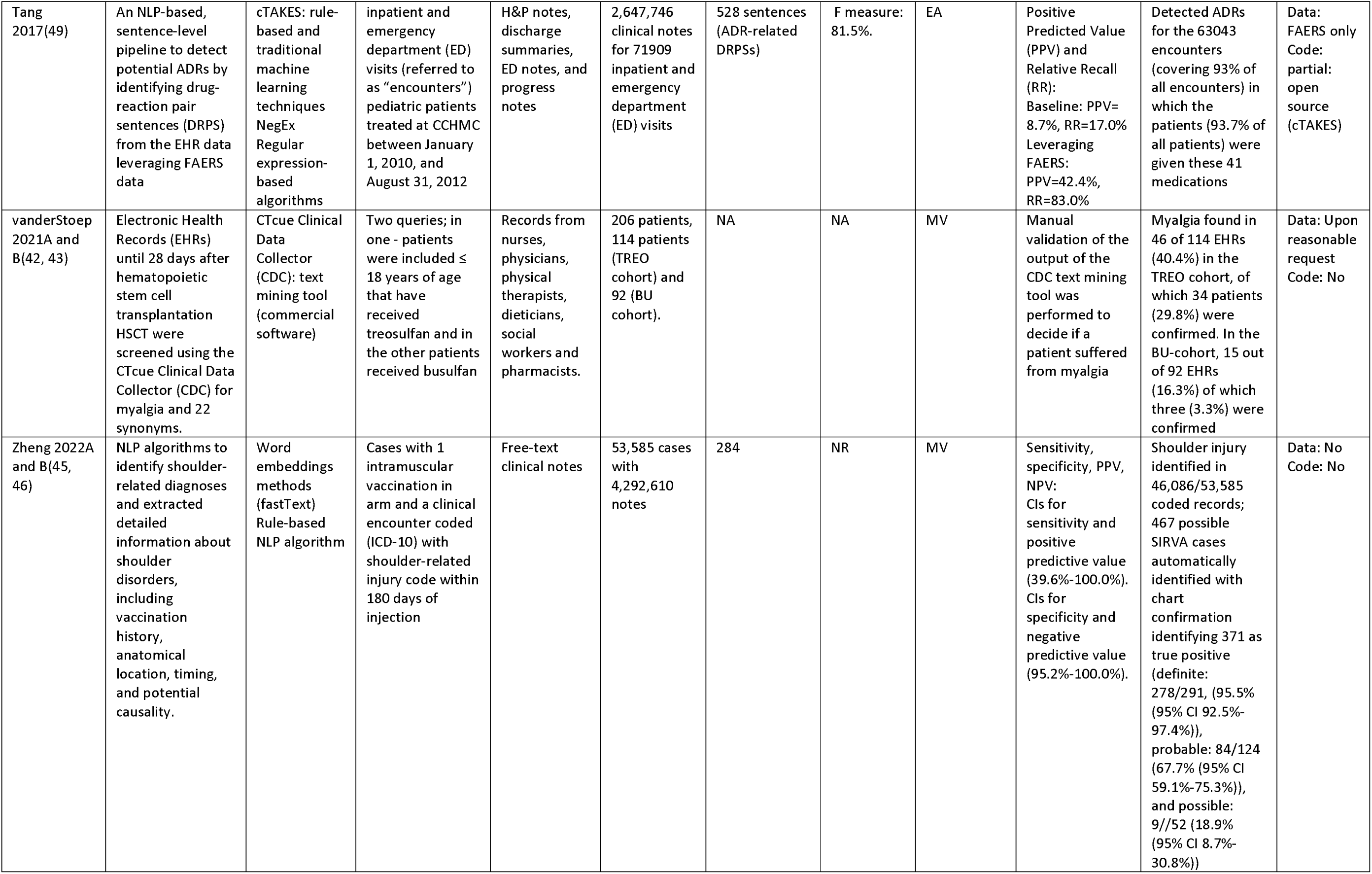
Methods and results of included studies.

While some studies described their dataset in terms of number of patients or number of unstructured notes, others reported both. In those that reported the number of notes used, the total varied from 32,000 to over four million.

The source data was not available in the studies, although three stated that it could be made available upon reasonable request (Matson 2023 (44), Miller 2022 (47, 48), vanderStoep 2021 (42, 43, 47, 48). Proprietary code developed, if any, was not made available, with three studies using open-source software (cTAKES).

### Conclusions within included studies

Extracting data from unstructured text may be particularly important in studying inequalities and co-morbidities, which may require large datasets. Findings from the studies are detailed in Table 4. Some studies emphasized the value of using EHR data to evaluate adverse events in children or by age group. For example, Ramsey 2021(39) demonstrated its value for studying a well-known adverse event in adults that had yet to be evaluated in children, Zheng 2022A and B (45, 46) differentiate between adult and pediatric populations, demonstrating that the rate of an adverse event can be different in the two groups, vanderStoep 2021 (42, 43) identified potential associations by age (<3 and >3) and Aldrich 2019 (38) looked at metabolizer status, which is well studied in adults, in children in relation to tolerability of SSRIs.

**Table 4:** Aims and conclusions of included studies.

| Study ID | Aim | Hypothesized benefit of clinical notes | Conclusion/Key Findings |
| --- | --- | --- | --- |
| Aldrich 2019(38) | To investigate the association between CYP2C19 metabolizer status and tolerability and treatment outcomes using ADEs extracted from the EHR. Comparison of EHR reported side effects were made with those reported in clinical trials. | NR | CYP2C19 metabolizer status helped to explain the wide variability in treatment outcomes. The number and type of side effects experienced was associated with metabolization rate, number of concomitant prescriptions, and adherence concerns. The frequency of side effects was higher than reported in prospective clinical trials. Collectively, our findings suggest that dosing es/citalopram based on CYP2C19 metabolizer status could improve safety and accelerate treatment response in pediatric patients. |
| Geva 2020(41) | To compare adverse drug event (ADE) rates determined from clinical notes using NLP to two RWD sources, electronic health records diagnostic codes and administrative claims data, among children treated with drugs for pulmonary hypertension. | Claims structured data for identifying diagnoses may lack sensitivity for detecting adverse drug events (ADEs), as not all signs and symptoms are recorded for billing purposes | Of 40 potential ADEs examined, 6 (15%) were identified significantly more frequently in the EHR clinical notes. An additional 13 potential ADEs were identified only in clinical notes but not in diagnostic codes. Compared to claims data, fourteen (35%) of 40 ADEs were found significantly more frequently in the free-text clinical notes. Analysis of clinical notes generally identifies more potential ADEs than diagnostic codes (a 7-fold increase) in either EHR or insurance claims datasets, but certain diagnoses are better represented in structured data. |
| Geva 2020(40) | To integrate a high-sensitivity natural language processing (NLP) pipeline for detecting potential adverse drug events (ADEs) with easily interpretable output for high-efficiency human review and adjudication of true ADEs | Manual review of notes to find relevant information is a time-consuming and laborious task, which has been made more difficult with the prevalence of copied text in EHR notes | ADEPT increased efficiency of this pharmacovigilance case study by reducing the time needed to review potential ADEs. ADEPT chart review typically took annotators less than 4 min per patient to complete, decreasing the time of chart review reported as 15–23 min in other studies. |
| Matson 2023(44) | To systematically assess the rate of adverse events of two-dose BNT162b2 vaccination in the pediatric population. High level comparison to previously published studies were reported. | NR | The reporting of adverse events remained low in passive surveillance. Serious adverse events were rare after the first and second doses of BNT162b2, with rates of anaphylaxis (six [0.01%] of 56 436), myocarditis (five [0.01%]), and pericarditis (three [0.01%]) consistent with previous studies. |
| Miller 2022A and B(47, 48) | To demonstrate the feasibility of developing an algorithm to ascertain a complex AE by capturing typhlitis using data from multiple EHR components. | Many clinically significant AEs are described primarily in clinician notes and radiology reports, we hypothesized that this novel approach of combining data from multiple EHR components, including discrete elements and those recorded as free text, would lead to accurate detection of typhlitis, and that automated identification would be more accurate than CRA-based and RN-based manually abstracted incidents of typhlitis in COG AE reports. | The automated algorithm identified true cases of typhlitis with higher sensitivity than Children's Oncology Group (COG) reporting. The algorithm identified false positives but reduced the number of courses needing manual review by 96% (961 to 37) by detecting potential typhlitis. This algorithm could provide a useful screening tool to reduce manual effort required for typhlitis AE reporting. |
| Ramsey 2019(39) | To evaluate the time course of es/citalopram-related weight gain in youth and the factors that influence its emergence using NLP to identify mentions in the EHR. | NR | The time course of documented weight gain was significantly associated with race ($p = 0.01$, log-rank test) but not by gender ($p = 0.58$). These findings highlight (1) the utility of using EMR-derived data to examine variability in side effects and (2) problems with “one size fits all” treatment monitoring strategies in youth and argue for precision medicine monitoring approaches |
| Tang 2017(49) | To determine whether the Food and Drug Administration’s Adverse Event Reporting System (FAERS) data set could serve as the basis to build an automated NLP-based, sentence-level pipeline to detect potential ADRs from electronic health record (EHR) monitoring for adverse drug events to reduce the cost of manual development of drug-ADR pairs and mitigate the need to chart review clinical notes in the EHR | A substantial portion of the meaningful information is represented only in clinical notes | Leveraging information from the FAERS reports, the performance of ADR detection was statistically significantly better than that of the baseline (manually curated drug-ADR pairs). The proposed semiautomated algorithm, when implemented in the clinical environments, could result in a substantial workload reduction for clinicians looking for ADRs in patients’ clinical notes. |
| vanderStoep 2021A and B(42, 43) | To investigate whether treosulfan compared with busulfan is associated with an increased risk of myalgia using a natural language processing and text-mining tool (CDC) to extract myalgia reports in the EHR. | The electronic health record (EHR) is an important source of data and contains valuable information collected during routine clinical practice, including side effects of drugs, however, this information is often stored in the EHR as free-text notes. Manual chart review is the gold standard for collection of data from EHRs, but this is laborious and very time-consuming. Natural language processing (NLP) and text mining techniques in the EHR can provide additional information about drugs that has not been discovered in clinical development. | Myalgia is a common adverse effect in treosulfan-based regimens in pediatric patients in the setting of HSCT, particularly in hemoglobinopathies. This study shows that retrospective studies can make an important contribution to the knowledge and recognition of adverse events. It provides valuable information, that can be included in the Summary of Product Characteristics of treosulfan. In the updated SmPC, pain in extremities is mentioned in the undesirable effects in the pediatric population with unknown frequency, this study found an incidence of 30%, predominantly in the hemoglobinopathy group. A text mining tool such as the CDC can help to detect adverse events more efficiently. |
| Zheng 2022A and B(45, 46) | To estimate the risk for shoulder conditions after vaccination and assess possible risk factors using NLP methods to extract cases of SIRVA from EHR notes. | Because there are no defined diagnosis codes for SIRVA, SIRVA case identification and determination must be done by reviewing free-text clinical documents. Manual review is both costly and time consuming; this challenge is magnified with SIRVA | NLP identified potential SIRVA cases in a small percentage of notes (0.9%), thus reducing the time spent for manual chart review for this rare event. These population-based data suggest a small absolute risk for shoulder conditions after vaccination. Given the high burden of shoulder conditions, clinicians should pay attention to any factors that may further increase risks. |

In addition to assessing the impact of age or adult versus pediatric groups, some studies looked at other characteristics. For example, Ramsey 2019 (39) and vanderStoep 2021A and B (42, 43) emphasize the value of EMR to accurately capture characteristics associated with an adverse event. In Ramsey 2021 (39) they evaluated differences by race and gender finding a significant difference between white and non-white patients, whereas vanderStoep 2021A and B (42, 43) identified potential associations with different co-morbidities.

There were also comparisons made with structured data or with other drugs. For example, Geva 2019 (41) found that more potential ADEs where found in clinical notes than when using diagnostic codes in either EHR or insurance claims datasets, while vanderStoep 2021A and B (42, 43) compares the rate of an adverse event for one drug to another drug.

Some studies were focused on the methods used and the benefits of automated detection of potential ADE mentions. For example, Tang 2017 (49) focused on the methodology of extracting data on multiple drugs and adverse events by leveraging FAERS reports to improve performance and again concluded that adverse event detection was possible. Miller 2022A and B (47, 48) and Zheng 2022A and B (45, 46) demonstrate the value of using EMR notes to identify rare and complex adverse events and potential time saving capabilities of their methods. Geva 2020 (40) developed a tool that combined the automated detection of potential ADEs with a user interface for validation, thus reducing the time spent in manual chart review.

## DISCUSSION

The included studies in this review demonstrate that unstructured data can be incorporated into pediatric pharmacovigilance systems as automatic mining can effectively and timely monitor adverse drug reaction (ADR) signals. The large volume of clinical notes included in the studies (over four million in one study), emphasises the power of NLP methods to scale studies as never before, and it is reasonable to expect that higher numbers could be processed with more recent advancements in NLP and processor technology. Extracting data from unstructured text may be particularly important in studying inequalities and co-morbidities which may require large datasets.

Using unstructured data in conjunction with structured data may help compensate for the disadvantages of each. Issues with unstructured data include incompleteness, inaccuracy and oversimplification (50). Structured data also suffers from no universally accepted set of coding algorithms or ICD codes for adverse drug events, making their consistent identification challenging (51). This has led to the development of automated methods with structured data (52, 53) and the creation of KidSIDES, a database of pediatric drug safety signals accessible through the PDSportal or as a bulk download (27).

The majority of the included studies used traditional NLP techniques, such as regular expressions and rule-based systems, to identify ADEs in unstructured clinical text. One of the main limitations of these methodologies is their lack of generalizability, relying heavily on predefined patterns and rules that must be meticulously crafted by experts (40, 54). Consequently, their effectiveness is tightly bound to the specific corpus they were designed for, often failing to generalize well across different ADEs and corpora. The reliance on exact matches also makes traditional NLP techniques prone to missing semantic meanings and struggle to interpret context, both crucial elements in unstructured clinical text.

Detecting ADEs in clinical text is challenging due to the complex nature of how these are documented. There is variable language used to describe ADRs, where different terms, non-standard descriptions, or abbreviations may be used to document the same reaction. Furthermore, medical mentions such as diagnoses, or signs and symptoms may be discussed as hypotheticals, e.g., discussions about potential ADEs that never occurred, or be documented as an absence of or a negated mention (55, 56).

The detection of ADEs in EHR data could greatly benefit from modern NLP techniques. The most recent study amongst the ones included, Matson 2023 (44), implemented a transformer-based classification model (57) to extract adverse events from EHR notes. The algorithm identified 18 different adverse event phenotypes associated with COVID-19 vaccination. In contrast with traditional techniques, the model was able to interpret the unstructured clinical text without relying on predefined patterns or rules, capturing nuanced language and semantic relationships more effectively.

However, one of the main drawbacks of transformer-based classification models is their requirement for extensive training, which involves significant manual effort. For instance, the algorithm described in Matson 2023 (44) was trained on 18,490 sentences manually labelled by clinical experts. This reliance on manual annotation represents substantial costs, with substantial financial resources required to compensate expert annotators and a lot of project time spent in the annotation process. This can be a limiting factor when applying these classification models to large-scale EHR data or expanding them to cover a wider range of ADEs.

Despite the prevalence of negated mentions in the EHR, only four studies reported identifying and excluding negated mentions as part of their methods (Geva A, Geva B, Miller, Tang), and only one (Miller) reported incorporating methods, such as ConText (58), to identify hypothetical or general discussions.

Non-standardized terminology and other documentation practices further complicate ADR detection. ADEs may be documented across multiple notes, by different providers, or in different sections of the clinical record. The practice of cutting and pasting forward portions of the clinical notes can lead to temporal ambiguity, which makes it difficult to determine if the ADE occurred before, during, or after medication administration (59–61). While the benefits of extracting potential ADEs were highlighted in the included studies, such as the reduction of time spent for manual chart review, the difficulty in accurately extracting a true ADE was made evident in the studies that validated their results. Finding ADEs automatically is challenging due to the multiple mentions of signs and symptoms in EHRs and linking these mentions to the drug as the cause or potential cause. Another challenge is the need for manual validation. In the included studies that validated the rate of true positives, it varied from <1% to 77% and this may be due to the way the source data was selected.

To address some of these challenges, researchers have recently explored the potential of large language models (LLMs). These advanced NLP algorithms can be effectively adapted to address specific tasks without the need for extensive training or manual data labelling, requiring only a reduced set of instructions in natural language (62).

LLMs are already being utilized to process unstructured medical data. Their capabilities in natural language understanding and their ability to follow clinical guidelines have been leveraged for matching patients to clinical trials (63) and supporting clinical decision-making (64). By incorporating domain-specific knowledge into the models, researchers are also employing LLMs to extract ADEs from unstructured clinical notes (65). These models have shown strong capabilities in understanding and interpreting the nuanced language typically found in EHRs, such as synonyms, abbreviations, and diverse semantic constructs used to describe ADEs. Moreover, LLMs’ ability to discern context enhances their utility in medical settings, allowing them to distinguish between actual medical events and hypothetical or negated mentions, a common challenge in traditional NLP methods when addressing the detection of ADEs from clinical notes. Consequently, we anticipate the potential of LLMs to significantly impact the analysis of ADEs in pediatric populations. This technology can enable the large-scale analysis of multiple adverse events, drastically reducing manual effort and surpassing the effectiveness of traditional NLP methods.

Another important aspect to consider is reproducibility. We found that none of the nine studies analyzed provided access to their developed code. Making the implementation of the developed NLP algorithms publicly available would enable other researchers to replicate the findings in different healthcare settings. Moreover, if accessible, these NLP methodologies could be adapted to study other ADEs of interest in pediatric populations.

The challenge of reproducibility is further compounded by limited data accessibility. Only three studies indicated data availability upon reasonable request - Matson 2023 (44) Miller 2022 (47, 48), vanderStoep 2021 (42, 43). This not only limits the ability to validate results but also hinders any meaningful comparative evaluation of different automated approaches to identify ADEs in EHR notes. One solution would be the creation of a benchmark dataset for this task. Indeed, there are several benchmark datasets that have been provided by the organizers of shared tasks, such as N2C2 (66) and MADE 1.0 (32). However, neither resource specifies the age of the included patients making it unclear whether pediatric populations were included in the annotated datasets.

Any release of annotated corpora, whether from a single study or benchmark datasets, requires the de-identification of protected health information (PHI) from the patient notes. De-identification of pediatric notes poses some specific considerations such as the mentions of family member’s names, occupation and medical history, mentions of schools and grade levels and growth or milestone mentions (67). Researchers should be aware of the special considerations needed for pediatric note de-identification and use tools that can be modified for this specific use case, such as Philter (68).

Among the analysed studies, only one, vanderStoep 2021A and B (42, 43), was conducted outside the United States, which limits the generalizability of findings to global pediatric populations. This predominance of studies from a single country emphasizes the need to diversify research efforts geographically to better understand the global variations in pediatric ADEs. Additionally, collecting datasets from multiple regions around the world will facilitate the development of more universally applicable ADE detection systems and ensure that pharmacovigilance tools can effectively serve pediatric populations globally.

The limited number of studies identified in this scoping review emphasises the lack of research in this field. It could be argued that as clinical trial evidence on adverse events can be limited and in particular adverse events in pediatric populations that research into other data sources for pediatric pharmacovigilance is more urgently required than for adults. However, while much of the literature focuses on the technical aspects of data mining of unstructured data few examples exist specifically for pediatrics. Some of the nine included studies in this review were from the same dataset. While the technical papers on methodologies for extraction from unstructured data may be generalizable from other age categories to pediatrics it is disappointing not to see more case examples of the application in pediatric populations where these methods may be most useful.

Others have argued that while sophisticated pipelines incorporating powerful NLP components have been developed, these approaches are rarely using in a “real” setting (69). A review of the use of NLP to analyse unstructured data in EHRs to identify patient-reported outcomes also found an emphasis on adult populations with only six of the 79 included studies in their systematic review focusing on pediatric populations (70). Yet it can be argued that filling the gaps in identifying ADEs in pediatric populations is more urgently required than for adult populations where more is already known.

In this review we have restricted the data source to unstructured clinical notes. However, there are other unstructured data sources such as social media that these methods may be applicable to. For example, two of our excluded studies, mined social media platforms one to detect signals between drugs and adverse events in preschool aged children with ADHD (71). Another study, mostly focused on adults, included suicidal thoughts or actions in children as an adverse event (72). Indeed, we have mined social media to extract data on birth defects in neonates (73) and other studies have explored using social media to detect adverse events in children (74, 75).

Another application for unstructured data in pediatric populations may be in studying adherence to documentation. Bannet 2024 used LLMs and has promising findings in monitoring documentation of side effects inquiry in clinic, telehealth or telephone consultations with pediatric patients. While this study does not aim to identify the adverse events themselves, it demonstrates another useful application of unstructured data in the field of pediatric adverse events monitoring – namely the adherence to documentation of adverse event inquiry.

### Strengths and Limitations

This is a fast-paced area of research: the applicability of our findings may change over time, particularly with advances in LLMs and AI.

While taking a comprehensive approach to searching for the included studies, few studies met our inclusion. There were also few comparisons to other data sources within these studies. This makes it difficult to fully assess the value of unstructured data in relation to structured data in electronic health records or to regulatory data or the scientific literature (including clinical trials and observational studies). Indeed, the most useful data may come from combining different data sources, but we were unable to validate this.

## CONCLUSIONS

This scoping review provides a valuable summary of research and important information for pharmacovigilance in children, as well as suggest future directions of further research in this area. Overall, the studies included demonstrated the effectiveness of NLP in identifying ADEs in children from clinical notes. However, the small number of included studies indicates that the use of unstructured data from clinical notes to monitor ADEs in children is not widely practiced.

## Data Availability

All data produced in the present work are contained in the manuscript

## SUMMARY POINTS

1. **High ADE Incidence in Pediatric Populations**: Children face heightened risks due to physiological differences and limited drug safety data specific to pediatrics. ADEs in children contribute to 10% of hospitalizations, with up to 45% being life-threatening.
2. **Off-Label Prescribing Risks**: The frequent off-label use of medications in pediatric care increases the risk of ADEs due to insufficient pediatric-specific data on drug efficacy and safety.
3. **Data Gaps and Challenges**: Limited pediatric-specific clinical trial data make it difficult to fully understand ADE patterns in children, leading to reliance on extrapolated adult data despite known differences in drug metabolism and responses.
4. **Limitations of Traditional Pharmacovigilance**: Current ADE surveillance systems, which rely on spontaneous reporting, suffer from underreporting and data incompleteness, limiting their effectiveness in detecting pediatric ADEs.
5. **Emerging Role of EHRs**: Electronic health records (EHRs) hold promise for enhancing pediatric pharmacovigilance, particularly through the use of natural language processing (NLP) to extract valuable data from unstructured clinical notes.
6. **Focus on Unstructured Data**: Despite its potential, research utilizing unstructured EHR data for detecting pediatric ADEs remains limited. Most studies to date focus on structured data or adult populations, highlighting the need for more pediatric-focused investigations.
7. **Future Directions**: Advances in NLP and machine learning, along with the integration of large datasets and diverse data sources, are expected to improve the detection and understanding of ADEs in pediatric populations, emphasizing the importance of ongoing research in this area.

## FUTURE ISSUES

1. **Improved Integration of Unstructured EHR Data with other data**: Combining unstructured electronic health record (EHR) data with other data sources could enhance the accuracy of ADE detection, but achieving integration remains challenging. Future work is required to develop tools that can overcome this.
2. **Utilization of Large Language Models (LLMs) for Pediatric ADEs**: LLMs offer potential for more efficient ADE detection in pediatric populations. However, the development of domain-specific LLMs that can perform with minimal training data and handle large-scale datasets remains an important area for future exploration.
3. **Ethical and Regulatory Considerations for Pediatric Pharmacovigilance**: The ethical challenges of pediatric pharmacovigilance, particularly regarding consent and data privacy, will require careful consideration. Developing ethical frameworks and regulations that support such research is critical and ways in which data sharing can be applied.
4. **Data Sharing for Systems Evaluation**: The lack of publicly available datasets prevents a comparative evaluation of the developed NLP methods. Albeit challenging due to deidentification, creating benchmark datasets and shared platforms could provide a foundation for rigorous evaluation and further advancement of systems designed to detect ADEs from clinical notes.
5. **Real-World Implementation**: Although sophisticated NLP methods have been developed, their real-world application in pediatric healthcare settings is limited. Future work should focus on practical implementation.
6. **Continuous Monitoring and Updating**: As medical knowledge and clinical practices evolve, so should the systems designed to detect ADEs in pedicatric populations. Ensuring these systems are continuously updated with new data and guidelines will be essential to maintain their relevance and effectiveness.
7. **Global Representation in Research:** Future research should prioritize conducting studies in diverse international settings to understand better the unique ADE profiles across different ethnicities and healthcare systems. Collaborative international research initiatives and partnerships can facilitate this requirement, helping to create a more inclusive and comprehensive approach to pediatric pharmacovigilance.

## SUPPLEMENTARY MATERIAL

**Supplementary Table 1:** Search databases searched with numbers of records retrieved.

| Database | Search Interface | Date Range | Date Searched | Records Retrieved |
| --- | --- | --- | --- | --- |
| MEDLINE | OVID | 1946 to September 04,2024 | 5<sup>th</sup> September 2024 | 184 |
| EMBASE | OVID | 1996 to 2024 Week 35 | 5<sup>th</sup> September 2024 | 307 |
| PsycINFO | OVID | 1987 to August 2024 Week 5 | 5<sup>th</sup> September 2024 | 25 |
| IEEE Xplore | ieeexplore-ieee-org | 2000-2025 | 5<sup>th</sup> September 2024 | 107 |
| ACM Digital Library | dl-acm-org | 2000-2025 | 5<sup>th</sup> September 2024 | 57 |
| Library, Information Science & Technology Abstracts (LISTA) | EBSCO | 2000-2025 | 5<sup>th</sup> September 2024 | 83 |
| Total |  |  |  | 763 |
| Total after duplicates removed |  |  |  | 636 |

Ovid MEDLINE(R) ALL <1946 to September 04, 2024>

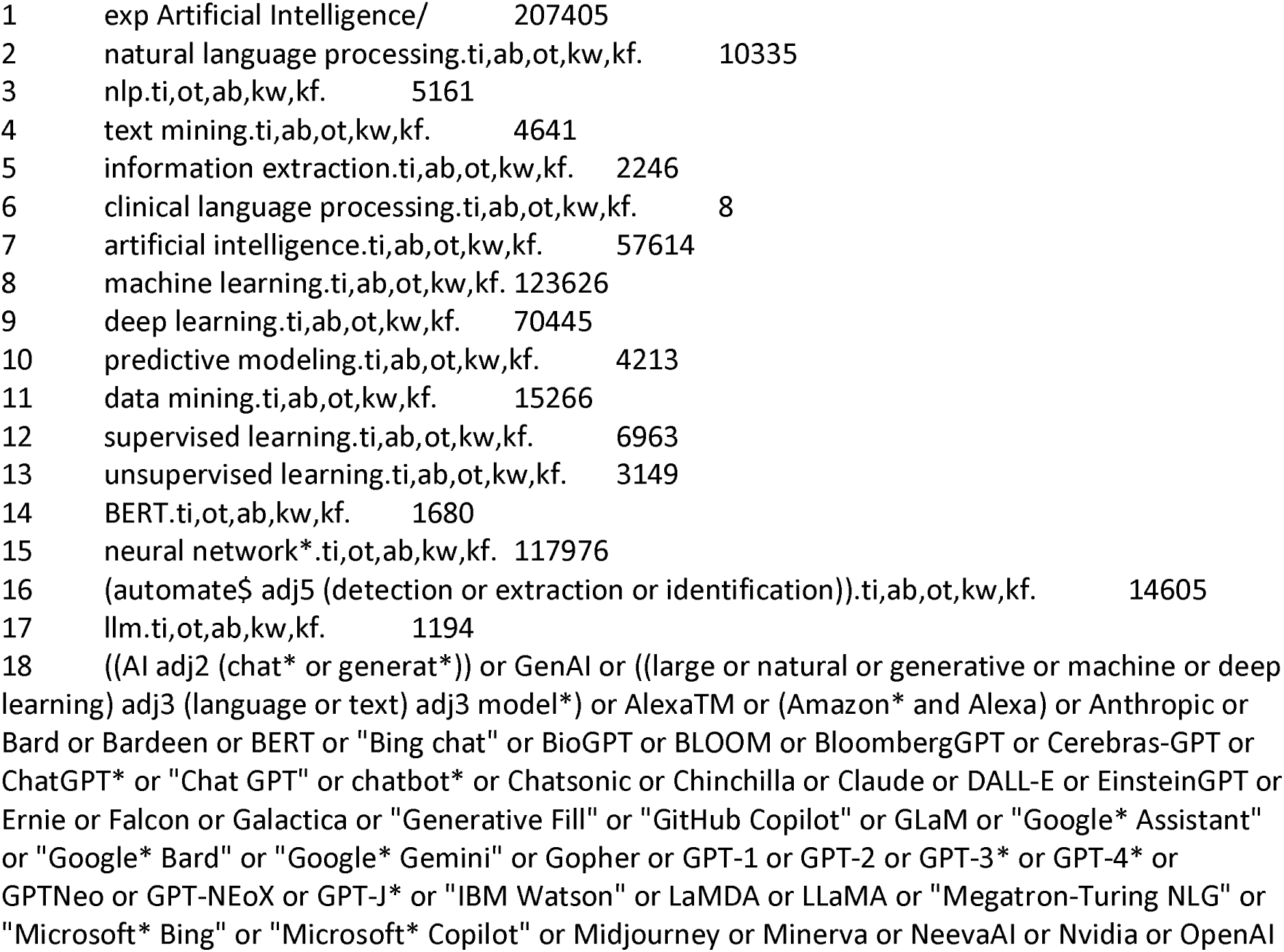
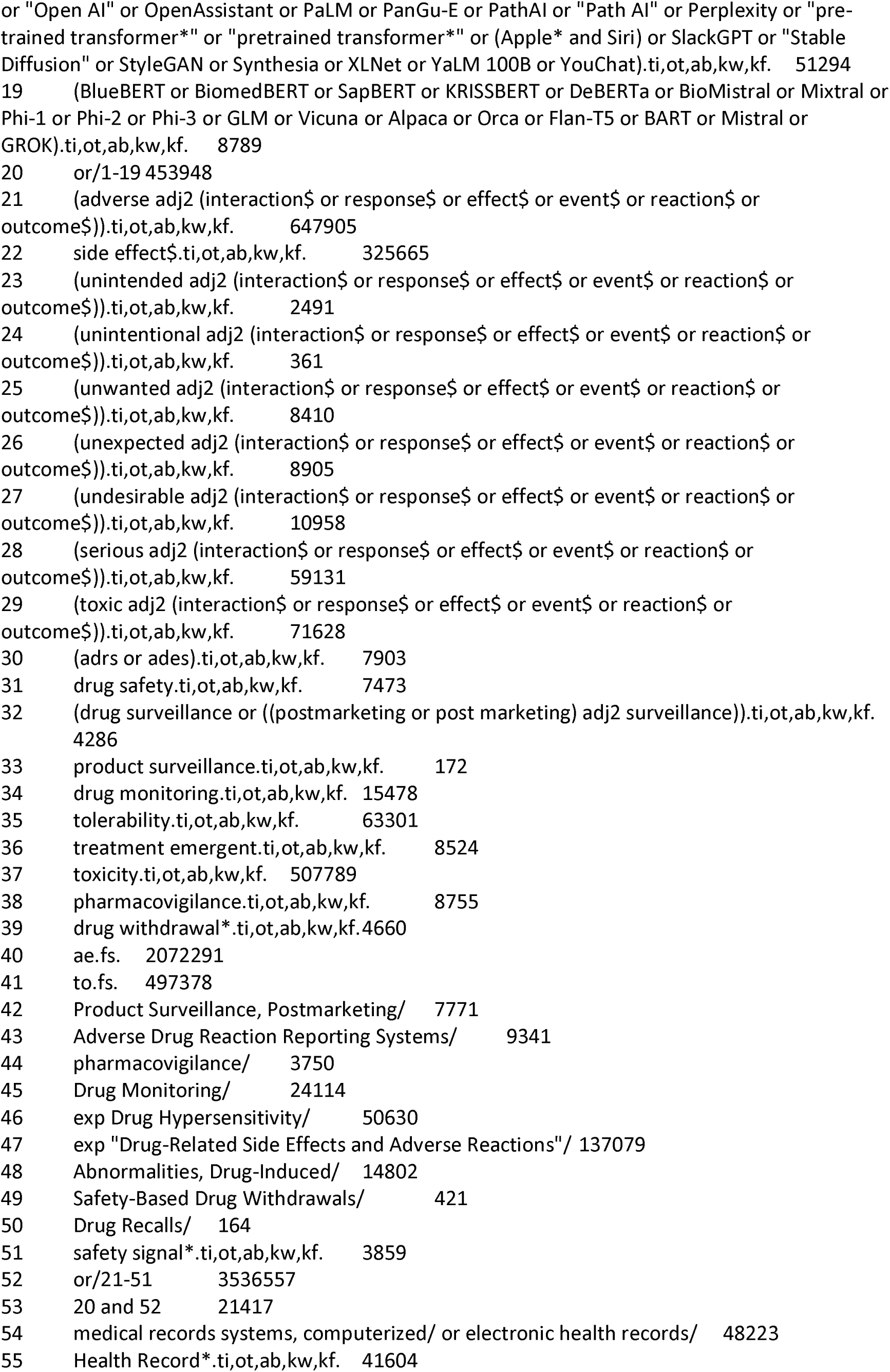
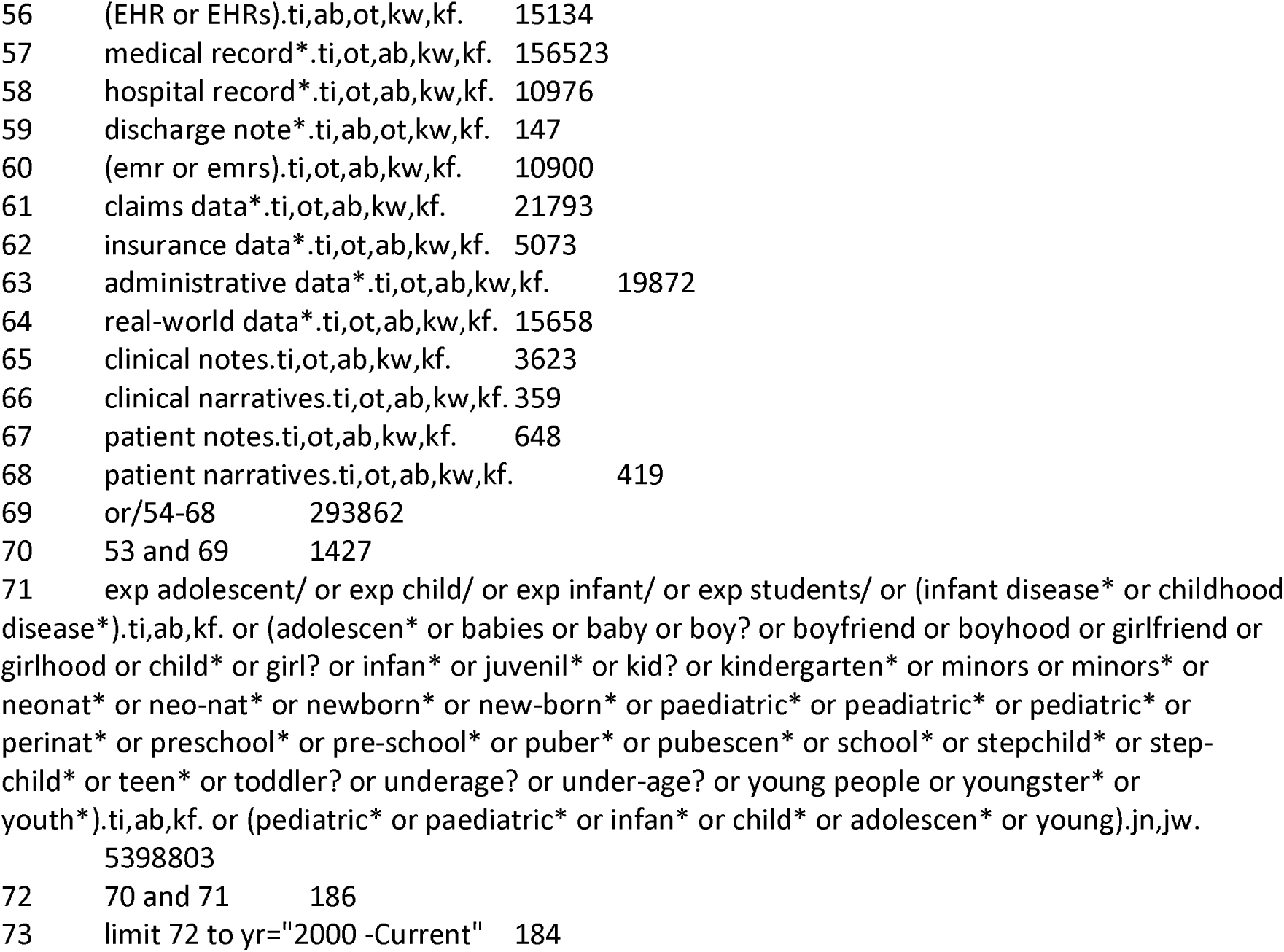

Embase <1996 to 2024 Week 35>

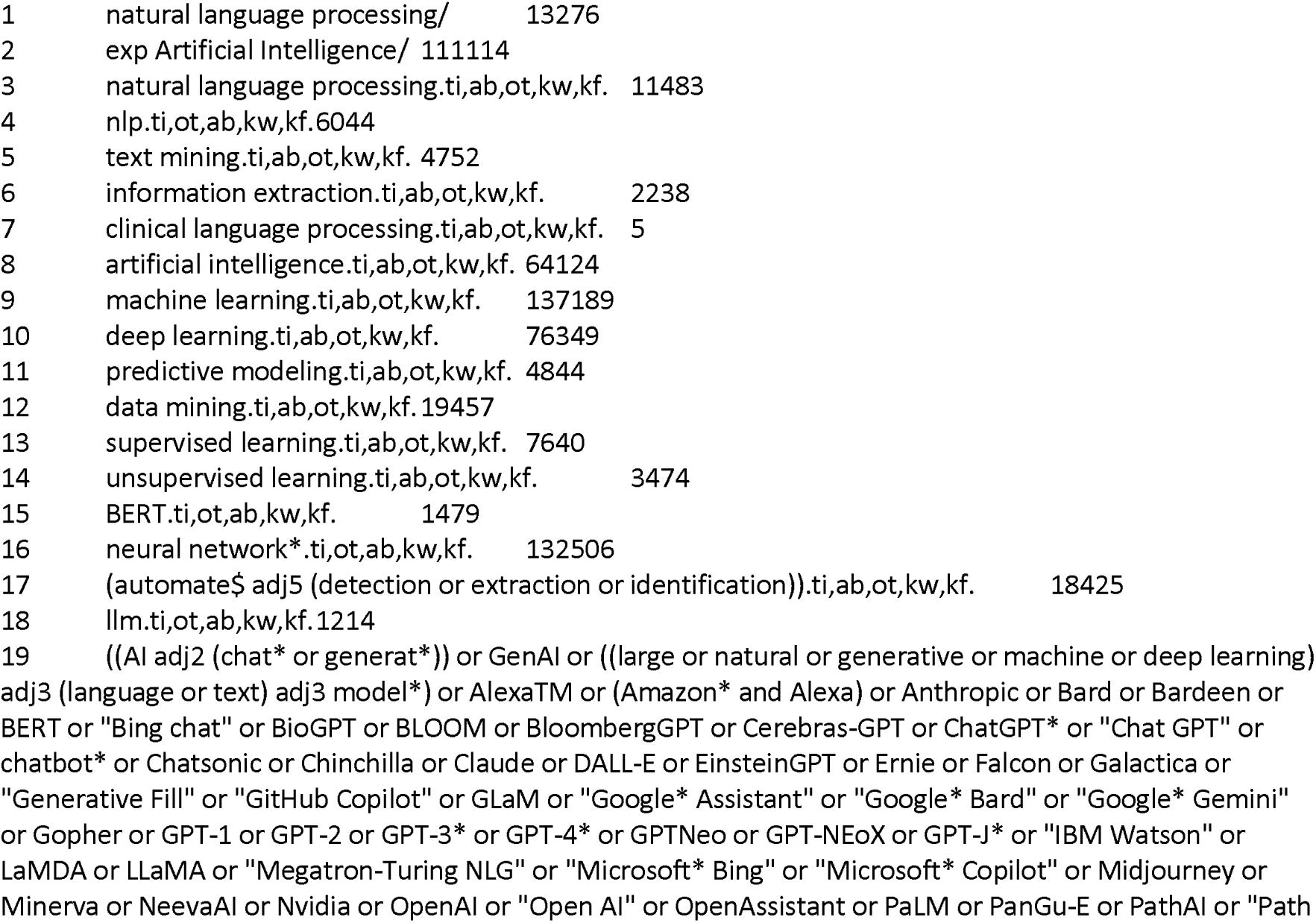
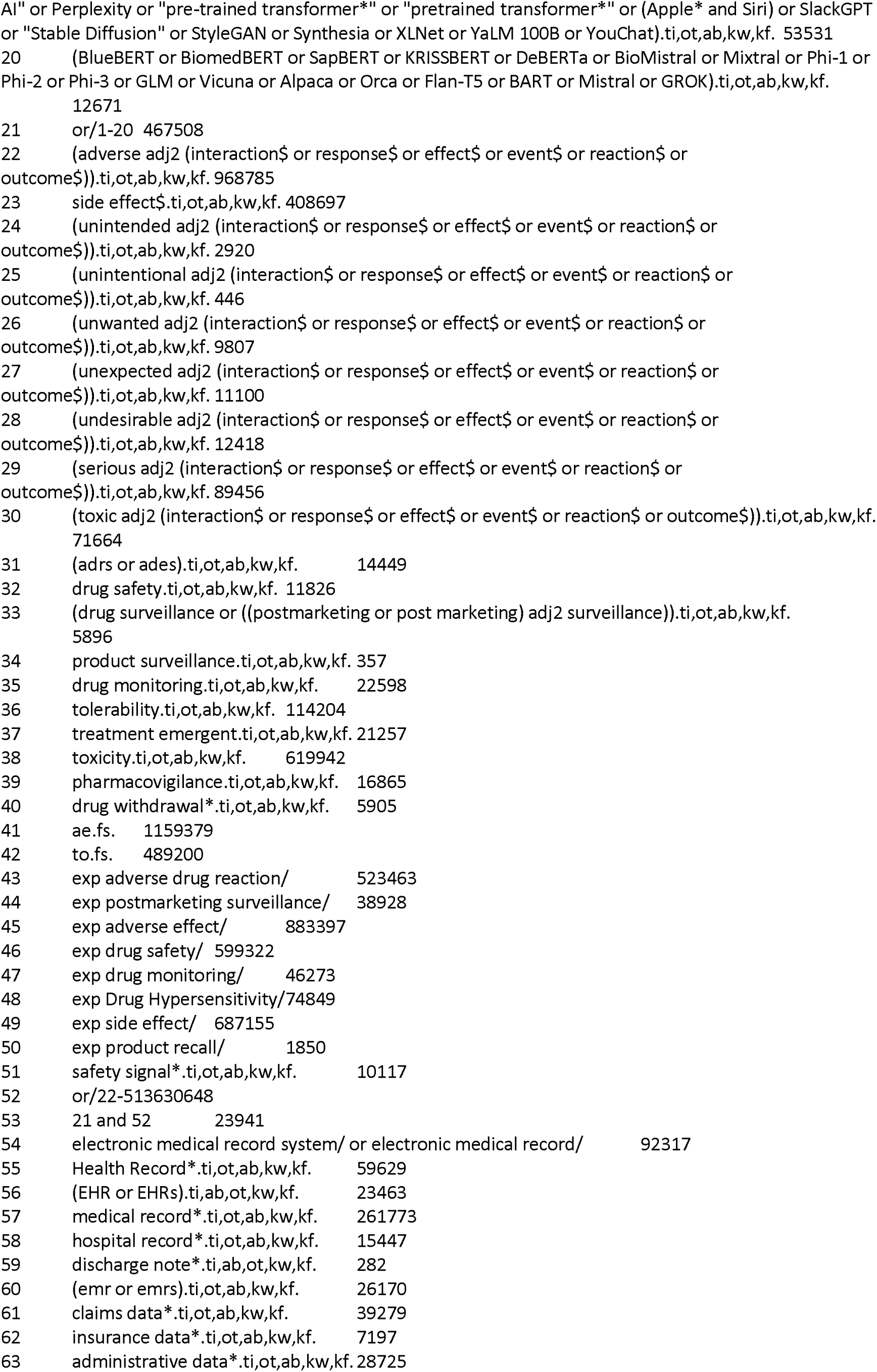
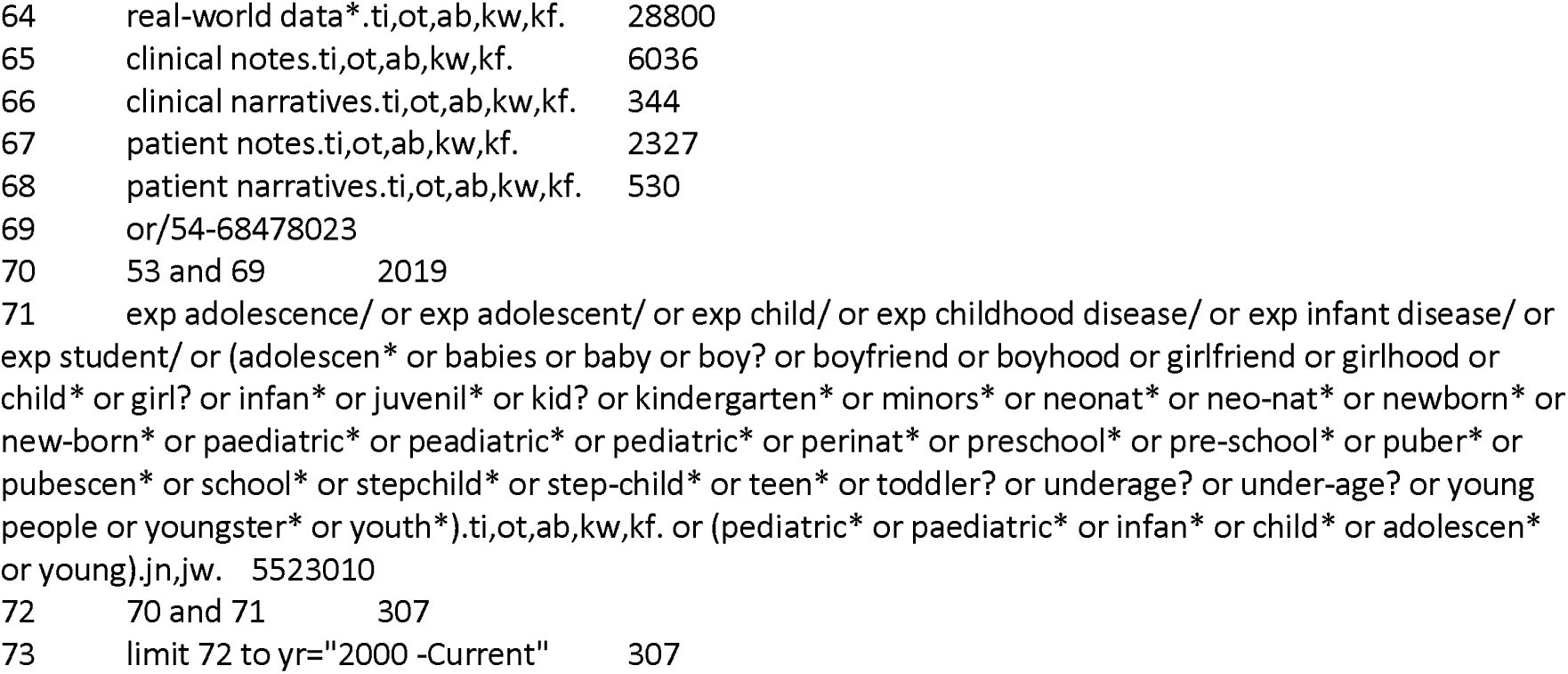

APA PsycInfo <1987 to August 2024 Week 5>

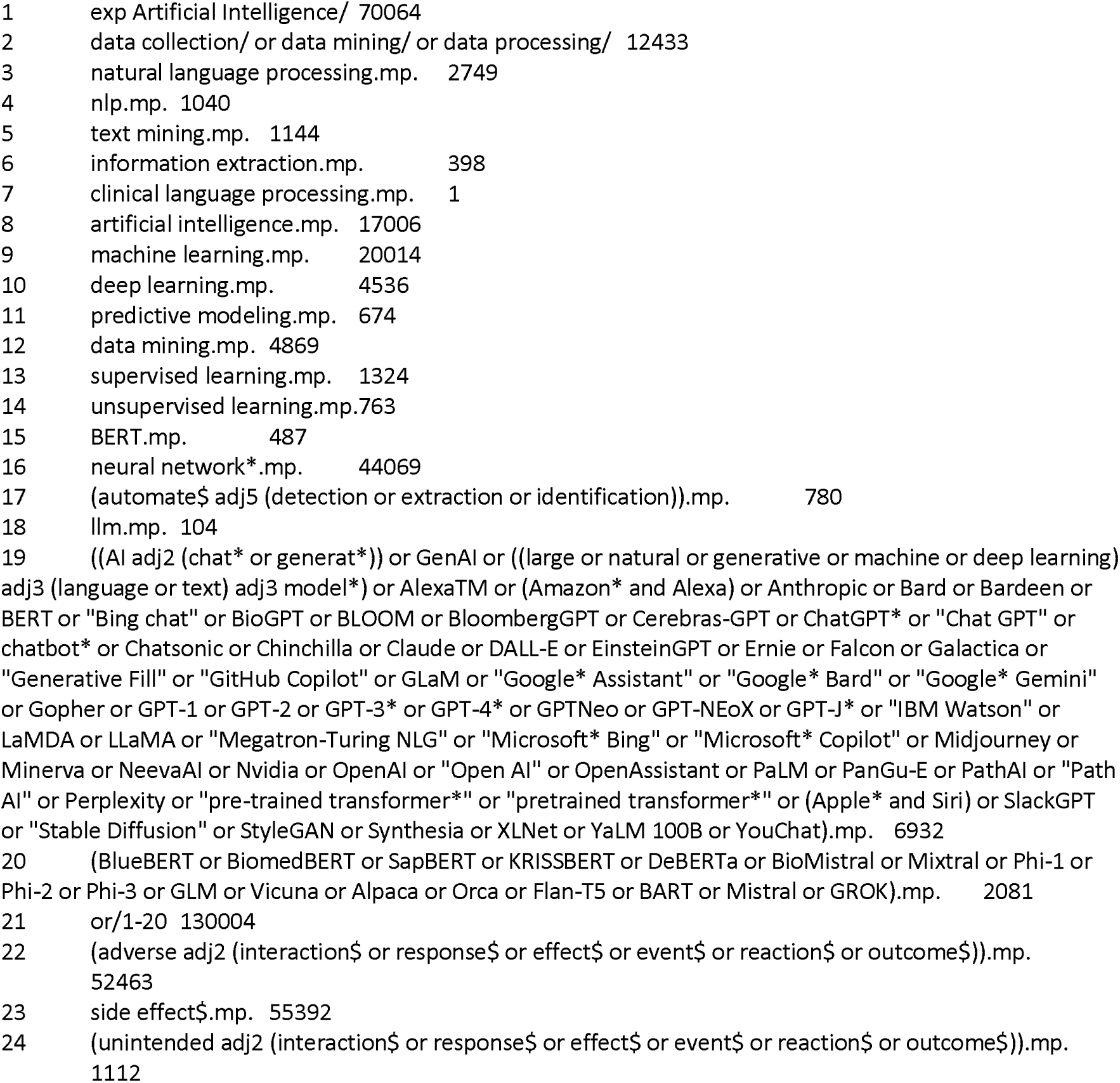
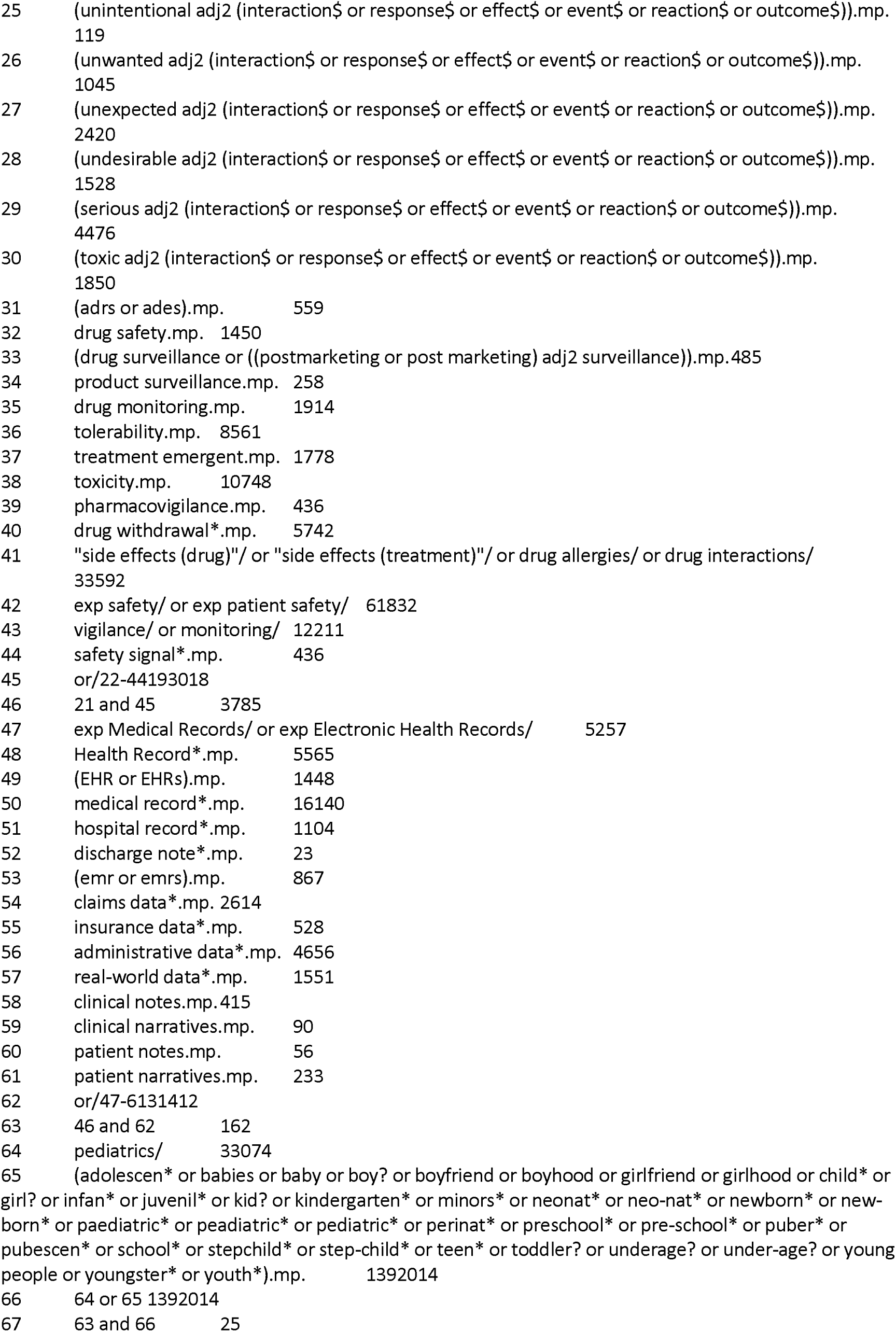

### 4. IEEE Xplore Search Strategy

All metadata:(adverse OR “side effect” OR “side effects” OR pharmacovigilance OR safety OR “drug surveillance” OR “drug monitoring” OR “post marketing surveillance” OR “postmarketing surveillance” OR “drug vigilance” OR toxicity OR “drug withdrawal” OR “drug withdrawals”) AND Title/Abstract: (“Health Record” OR “Health Records” OR “medical record” OR “medical records” OR “hospital record” OR “hospital records” OR “discharge note” OR “discharge notes” OR “clinical notes” OR “claims data*” OR “administrative data*” OR “insurance data*” OR “real-world data*” OR “clinical narratives” OR “patient notes” OR “patient narratives”) AND All metadata:(newborn OR “new-born” OR child* OR infant* OR pediatric* OR paediatric* OR adolescen* OR teenagers OR “young people” OR youth)

Limit year 2000 to 2024

107 records

### 5. ACM Digital Library Search Strategy

Two stage search due to limitations of the search interface:

Search 1

Title: (adverse OR “side effect*” OR pharmacovigilance OR safety OR “drug surveillance” OR “drug monitoring” OR “post marketing surveillance” OR “postmarketing surveillance” OR “drug vigilance” OR toxicity OR “drug withdrawal” OR “drug withdrawals”)

Anywhere: (“Health Record” OR “Health Records” OR “medical record” OR “medical records” OR “hospital record” OR “hospital records” OR “discharge note” OR “discharge notes” OR “clinical notes” OR “claims data” OR “administrative data” OR “insurance data” OR “real-world data” OR “claims database” OR “administrative database” OR “insurance database” OR “real-world database” OR “claims databases” OR “administrative databases” OR “insurance databases” OR “real-world databases” OR “clinical narratives” OR “patient notes” OR “patient narratives”)

Anywhere: (newborn OR “new-born” OR child* OR infant* OR pediatric* OR paediatric* OR adolescen* OR teenagers OR “young people” OR youth)

Results: 22 records

Search 2

Anywhere: (adverse OR “side effect*” OR pharmacovigilance OR safety OR “drug surveillance” OR “drug monitoring” OR “post marketing surveillance” OR “postmarketing surveillance” OR “drug vigilance” OR toxicity OR “drug withdrawal” OR “drug withdrawals”)

Title: (“Health Record” OR “Health Records” OR “medical record” OR “medical records” OR “hospital record” OR “hospital records” OR “discharge note” OR “discharge notes” OR “clinical notes” OR “claims data” OR “administrative data” OR “insurance data” OR “real-world data” OR “claims database” OR “administrative database” OR “insurance database” OR “real-world database” OR “claims databases” OR “administrative databases” OR “insurance databases” OR “real-world databases” OR “clinical narratives” OR “patient notes” OR “patient narratives”)

Anywhere: (newborn OR “new-born” OR child* OR infant* OR pediatric* OR paediatric* OR adolescen* OR teenagers OR “young people” OR youth)

Results: 36 records

#### LISTA

All text:(adverse OR “side effect*” OR pharmacovigilance OR safety OR “drug surveillance” OR “drug monitoring” OR “post marketing surveillance” OR “postmarketing surveillance” OR “drug vigilance” OR toxicity OR “drug withdrawal*”) AND

All text: (“Health Record*” OR “medical record*” OR “hospital record*” OR “discharge note*” OR “clinical notes” OR “claims data*” OR “administrative data*” OR “insurance data*” OR “real-world data*” OR “clinical narratives” OR “patient notes” OR “patient narratives”) AND

All text:(newborn OR “new-born” OR child* OR infant* OR pediatric* OR paediatric* OR adolescen* OR teenagers OR “young people” OR youth)

Results: 83 records

## EXCLUDED STUDIES

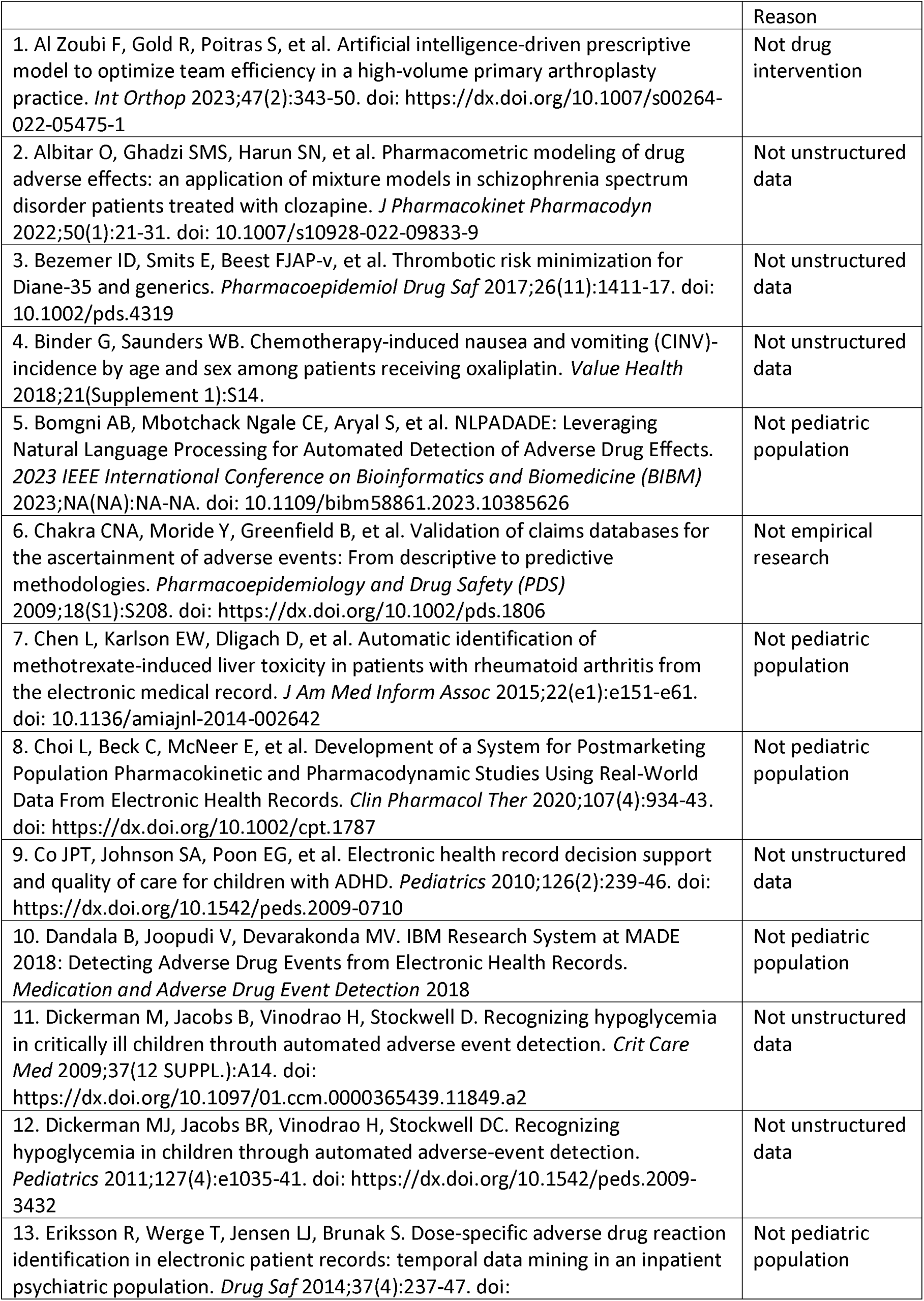
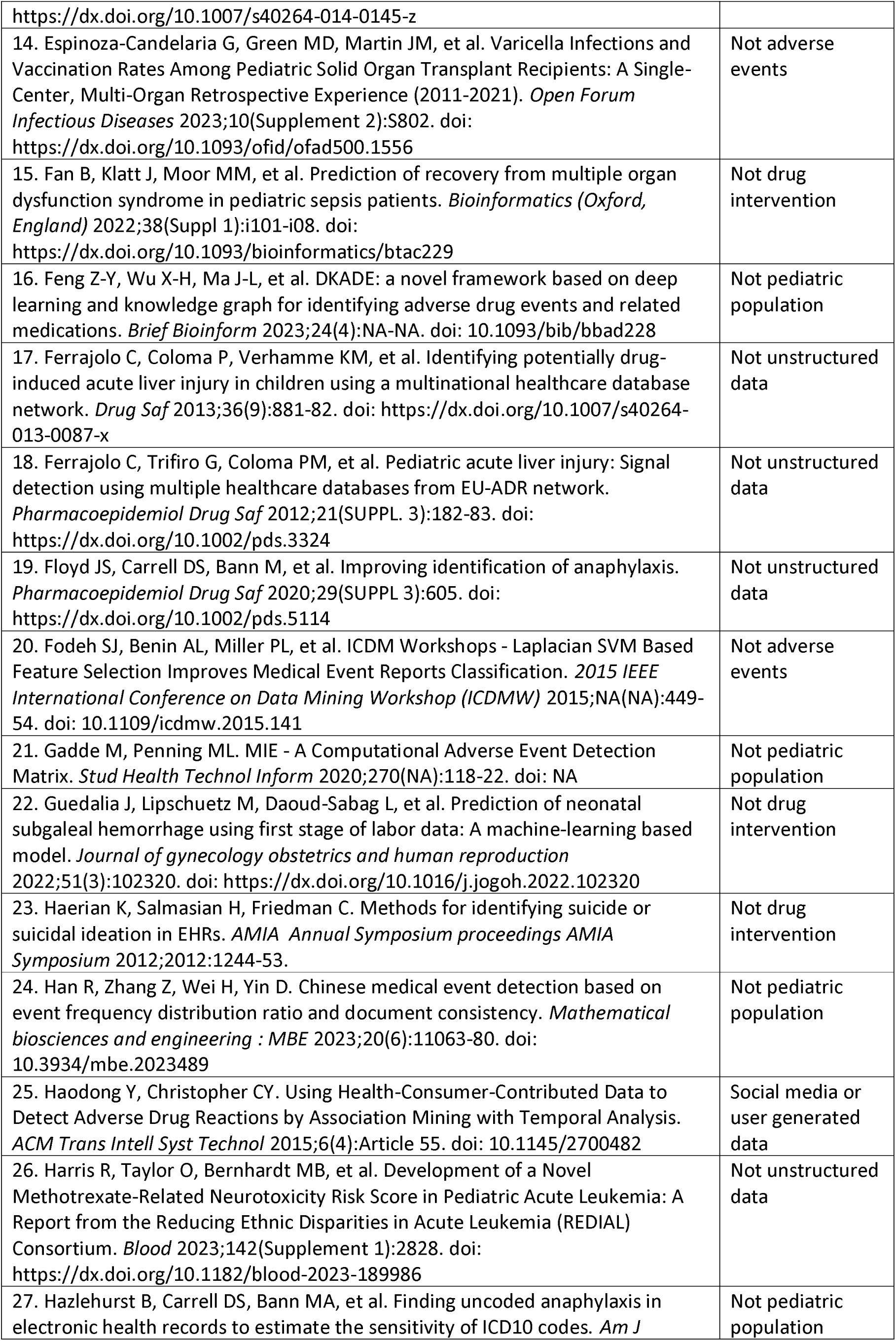
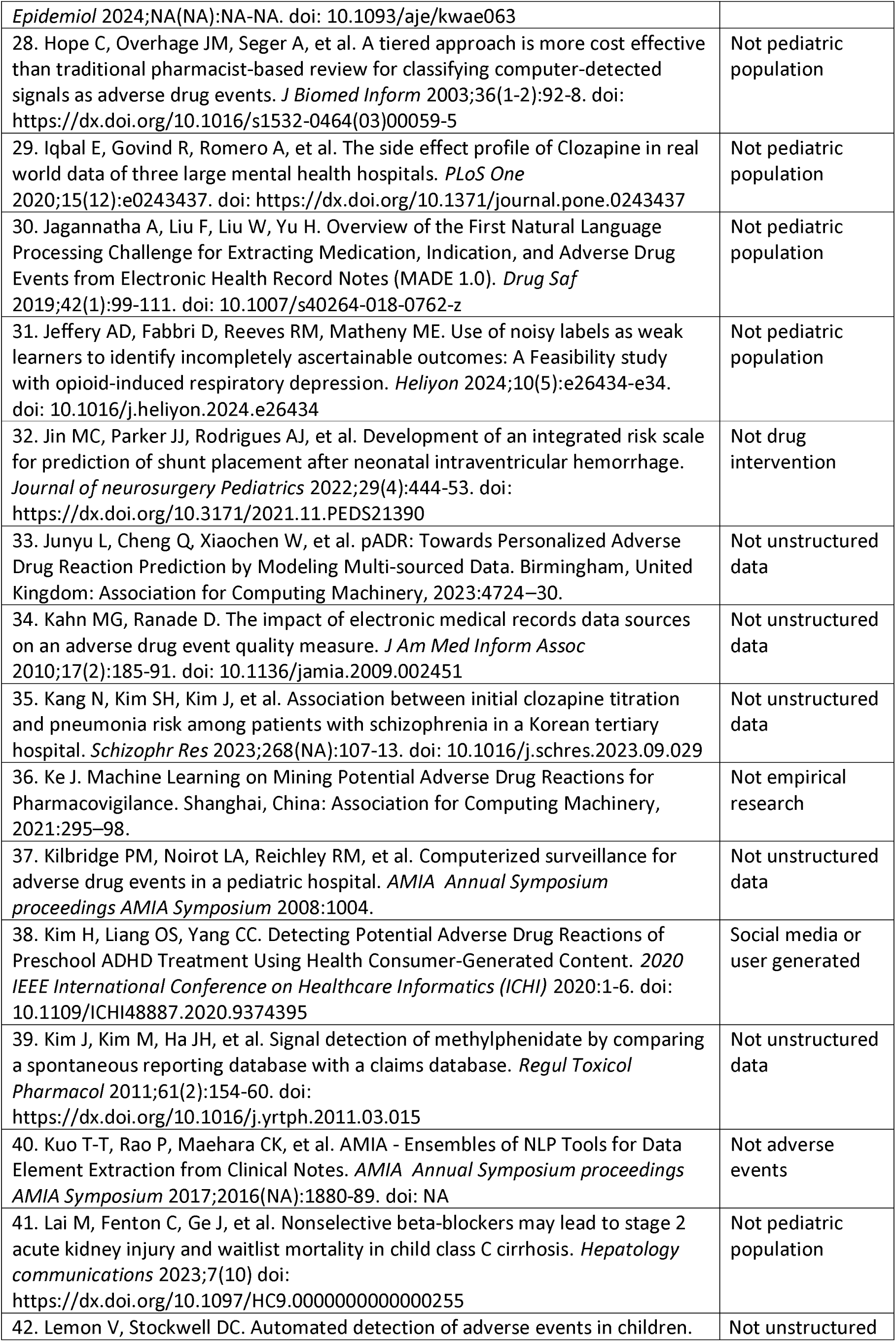
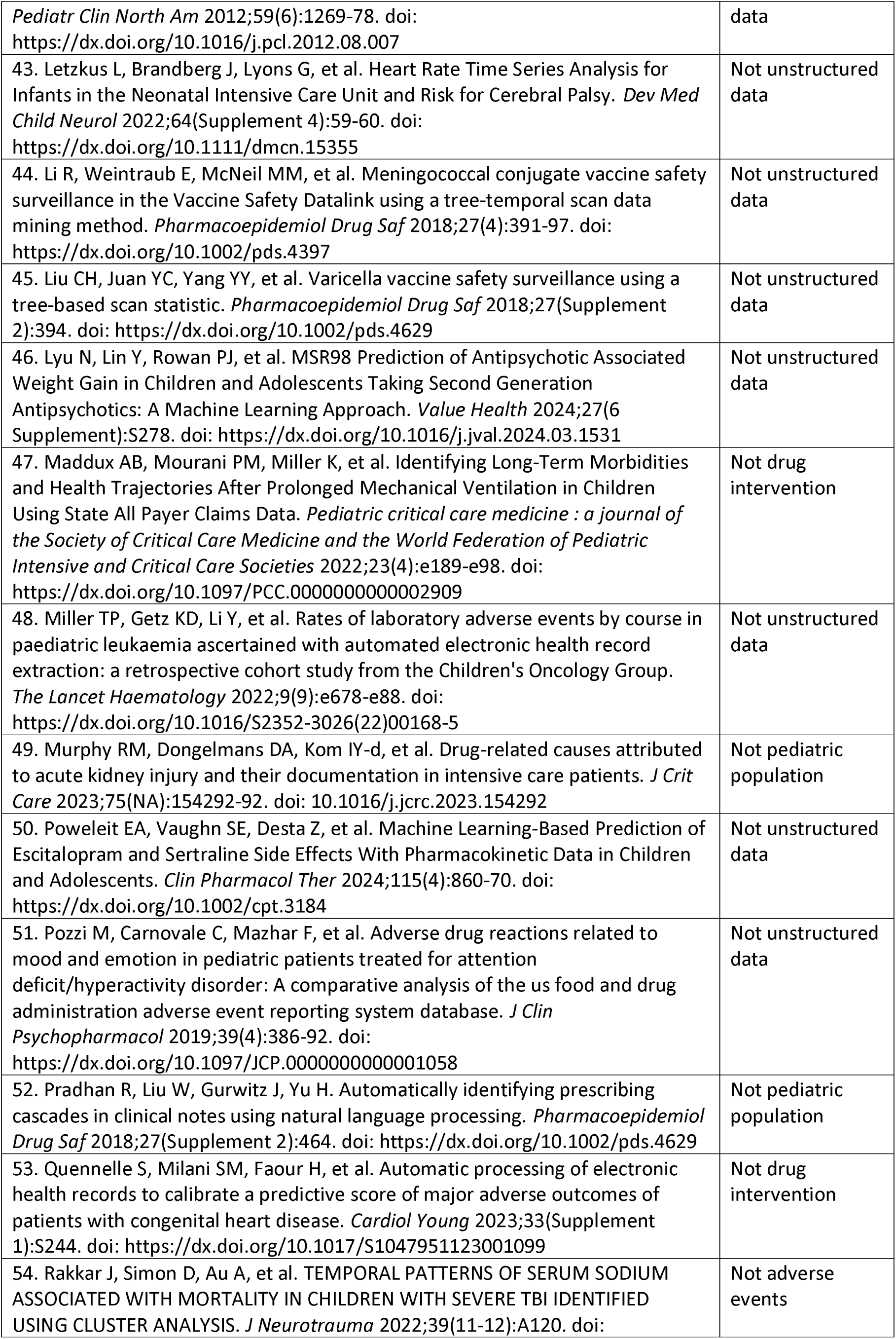
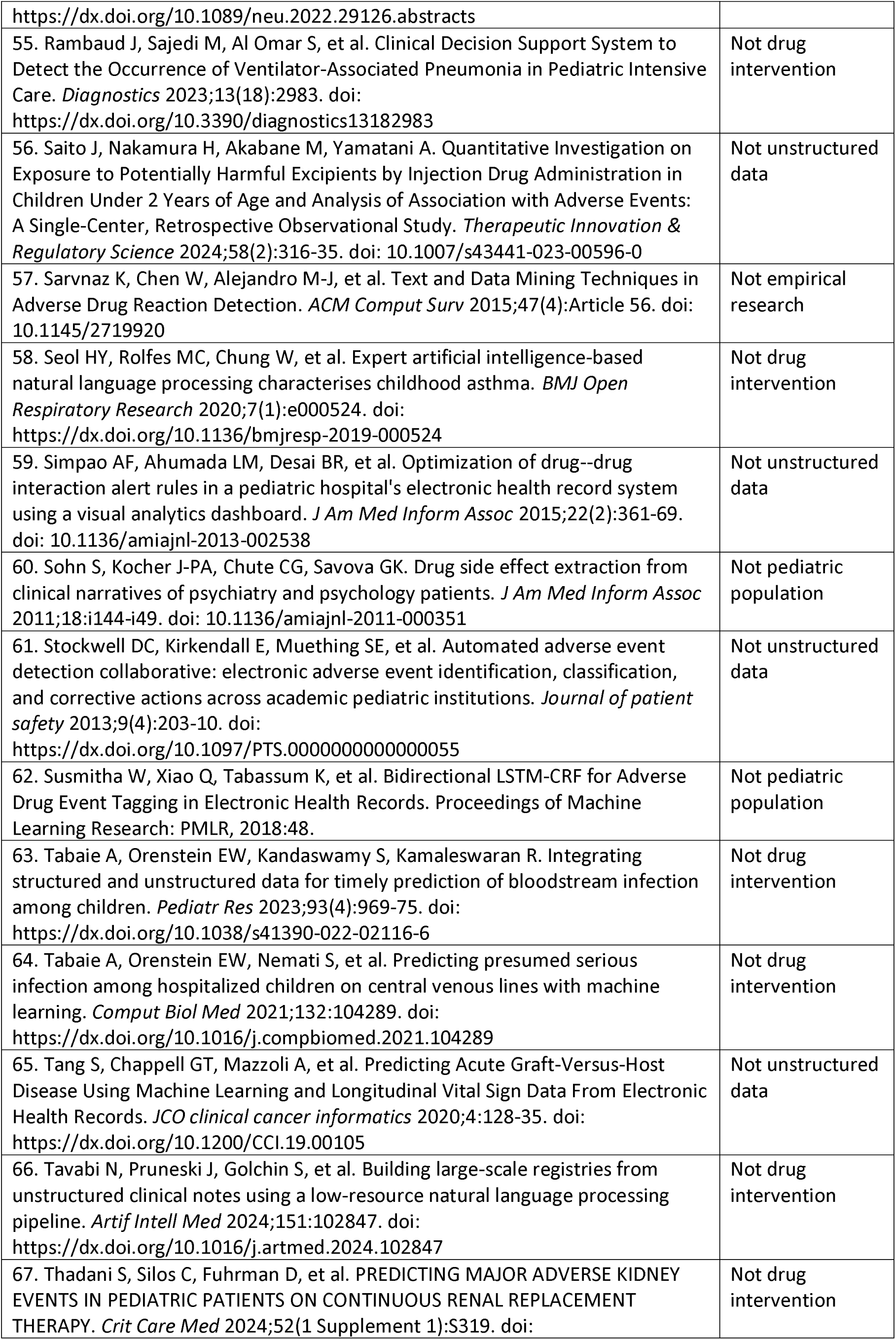
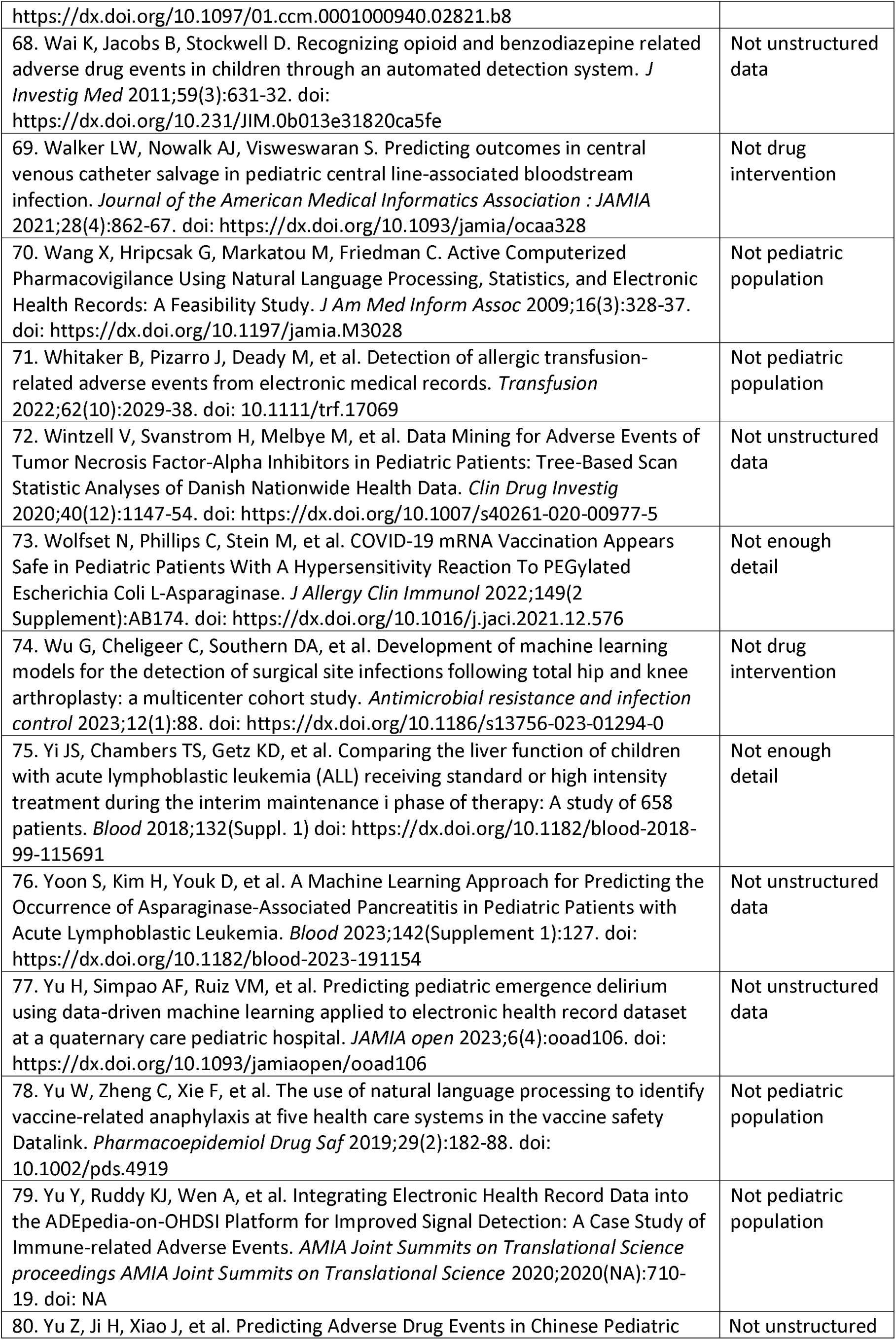
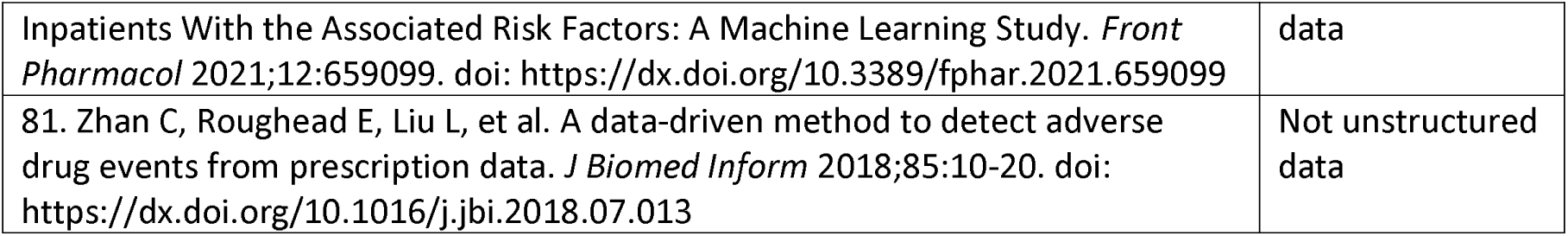

